# Excitation-inhibition imbalance in Alzheimer’s disease using multiscale neural model inversion of resting-state fMRI

**DOI:** 10.1101/2022.10.04.22280681

**Authors:** Guoshi Li, Li-Ming Hsu, Ye Wu, Andrea C Bozoki, Yen-Yu Ian Shih, Pew-Thian Yap

**Affiliations:** Department of Radiology University of North Carolina Chapel Hill, NC USA; Biomedical Research Imaging Center University of North Carolina Chapel Hill, NC USA; Center for Animal MRI University of North Carolina Chapel Hill, NC, USA; Department of Neurology University of North Carolina Chapel Hill, NC, USA

## Abstract

Alzheimer’s disease (AD) is a serious neurodegenerative disorder without a clear understanding of the etiology and pathophysiology. Recent experimental data has suggested neuronal excitation-inhibition (E-I) imbalance as an essential element and critical regulator of AD pathology, but E-I imbalance has not been systematically mapped out for either local or large-scale neuronal circuits in AD. By applying a Multiscale Neural Model Inversion (MNMI) framework to the resting-state functional MRI (rs-fMRI) data from the Alzheimer’s Disease Neuroimaging Initiative (ADNI), we identified brain regions with disrupted E-I balance based on impaired mesoscale excitatory and inhibitory connection strengths in a large network during AD progression. We observed that both intra-regional and inter-regional E-I balance is progressively disrupted from cognitively normal individuals, to mild cognitive impairment (MCI) and to AD, and E-I difference (or ratio) can be abnormally increased or decreased, depending on specific region. Also, we found that (local) inhibitory connections are more significantly impaired than excitatory ones and the strengths of the majority of connections are reduced in MCI and AD, leading to gradual decoupling of neural populations. Moreover, we revealed a core AD network comprised mainly of limbic and cingulate regions including the hippocampus, pallidum, putamen, nucleus accumbens, inferior temporal cortex and caudal anterior cingulate cortex (cACC). These brain regions exhibit consistent and stable E-I alterations across MCI and AD, and thus may represent early AD biomarkers and important therapeutic targets. Lastly, the E-I difference (or ratio) of multiple brain regions (precuneus, posterior cingulate cortex, pallium, cACC, putamen and hippocampus) was found to be significantly correlated with the Mini-Mental State Examination (MMSE) score, indicating that the degree of E-I impairment is behaviorally related to MCI/AD cognitive performance. Overall, our study constitutes the first attempt to delineate E-I imbalance in large-scale neuronal circuits during AD progression, which may facilitate the development of new treatment paradigms to restore pathological E-I balance in AD.

## Introduction

Alzheimer’s disease (AD) is neurodegenerative disorder characterized by progressive and irreversible cognitive decline (Bateman et al., 2012). It is the leading cause of dementia affecting more than 47 million people worldwide and this number is expected to increase to 131 million by 2050 (Tiwari et al., 2019). The healthcare cost for patients with AD and other dementias is enormous and is estimated to be 236 billion in the US for 2016 alone and predicted to quadruple by 2050 (Alzheimer’s Association, 2016). Despite decades of extensive research, a clear understanding of the etiology and pathophysiology of AD remains elusive. Current treatments are only symptomatic without slowing down the progression of the disease (Aldehri et al., 2018). The lack of effective treatment highlights the paramount importance of identifying new pathophysiological and therapeutic targets (Thakur et al., 2018).

Excitation-inhibition (E-I) balance represents a promising pathophysiological and therapeutic target for AD. <u>First</u>, disrupted E-I balance may underlie the key pathophysiological mechanism of AD. One of the pathological hallmarks of AD is the accumulation of amyloid-β (Aβ) peptides in the brain that occurs long before clinical disease onset (Karran et al., 2011; Huang and Mucke, 2012). During this long extended preclinical stage, soluble Aβ oligomers and amyloid plaques disrupt neuronal circuit activity and function by altering synaptic transmission and E-I balance leading to cognitive malfunction (Palop and Mucke, 2010; Busche and Konnerth, 2016; Palop and Mucke, 2016). In particular, high Aβ levels elicit epileptiform discharges and non-convulsive seizures in both hippocampal and neocortical networks of human amyloid precursor protein (hAPP) transgenic mice (Palop et al., 2007), which closely relates to the increased incidence of epileptic seizures in AD patients (Palop and Mucke, 2009). <u>Second</u>, E-I disruption is not only the consequence of Aβ deposit, but also a driver of the amyloid pathology. Experimental data indicate that Aβ release is regulated by neuronal activity (Nitsch et al., 1993; Bero et al., 2011) and driven by increased metabolism (Cohen et al., 2009; Johnson et al., 2014). Also, Aβ accumulation is associated with enhanced neural activity in task-related regions during memory encoding (Mormino et al., 2012) and reduction of neural hyperactivity decreases Aβ aggregation as well as axonal dystrophy and synaptic loss (Yuan and Grutzendler, 2016). <u>Lastly</u>, restoration of E-I balance has been shown to rescue circuit dysfunction and ameliorate cognitive impairments in both AD mouse models (Verret et al., 2012; Busche et al., 2015; Yuan and Grutzendler, 2016) and humans with early AD (Bakker et al., 2012), suggesting a direct link between E-I imbalance and cognitive malfunction. Taken together, these findings emphasize the significance of identifying E-I imbalance in AD, particularly in the initial disease stage for early diagnosis and intervention.

Functional magnetic resonance imaging (fMRI) is a core noninvasive method to measure brain activity (Glover, 2011) and has been widely used to study functional network alterations in AD (e.g. Filippi and Agosta, 2011; Brier et al., 2014; Dennis and Thompson, 2014). These studies have revealed both abnormal brain network activation/deactivation and dysfunctional connectivity patterns in AD involving the default mode (DMN), salience, executive control and limbic networks (Lustig et al., 2003; Dickerson et al., 2004, 2005; Celone et al., 2006; Greicius et al., 2004; Menon, 2011; Dhanjal and Wise, 2014; Badhwar et al., 2017; Schultz et al., 2017). However, conventional fMRI cannot distinguish E and I activity because fMRI signal increases regardless of selective E or I activation (Devor et al., 2007; Anenberg et al., 2015; Vazquez et al., 2018). This is not surprising, as activation of inhibitory neurons also consumes energy and triggers subsequent vascular signaling cascades that drive functional hyperemia (Anenberg et al., 2015; Uhlirova et al., 2016; Vazquez et al., 2018). Moreover, most current analytic approaches for fMRI, including graph theory, seed-based approaches, and independent component analysis (Li et al., 2009; Sporns, 2014; Preti et al., 2017) do not allow for determination of causal relationships between regions, nor do they provide insight into the dynamic meso-scale neuronal relationships that underpin blood-oxygen-level-dependent (BOLD) signal variations, thus unable to identify E-I imbalance at circuit levels. Generative modeling, by comparison, builds on biologically plausible models of neural interactions (Friston et al., 2003; Friston, 2011; Stephan et al., 2015; Li and Yap, 2022) and thus can, in principle, resolve excitatory versus inhibitory neuronal activity. For example, de Hann et al. (2012, 2017) developed a large-scale neural mass model to examine the effects of excessive neuronal activity on functional network topology and dynamics. In the first study (de Hann et al., 2012), they demonstrated that synaptic degeneration induced by neuronal hyperactivity results in hub vulnerability in AD including loss of spectral power and long-range synchronization. In a subsequent study (de Hann et al., 2017), paradoxically, the authors found that selective stimulation of all excitatory neurons in the network leads to sustained preservation of network integrity in the presence of activity-dependent synaptic degeneration. Using a computational framework termed “The Virtual Brain (TVB)”, Zimmerman et al., (2018) estimated personalized local excitation and inhibition parameters as well as global coupling strength based on resting-state fMRI (rs-fMRI) data from healthy individuals and patients with amnestic MCI (aMCI) and AD. They demonstrated that the model parameters required to accurately simulate empirical functional connectivity (FC) significantly correlate with cognitive performance, which surpasses the predictive capability of empirical connectomes. More recently, van Nifterick et al. (2022) proposed a multiscale brain network model to link AD-mediated neuronal hyperactivity to large-scale oscillatory slowing observed from magnetoencephalography (MEG) data in human early-stage AD patients. They modified relevant model parameters to simulate six literature-based cellular conditions of AD and compared them to healthy and non-AD scenarios. It was found that neuronal hyperactivity can indeed result in oscillatory slowing via either over-excitation of pyramidal cells or decreased excitability of inhibitory interneurons, supporting the hypothesis that E-I imbalance underlies whole-brain network dysfunction in prodromal AD. Nevertheless, all these previous studies focused on cellular/network simulation or AD differentiation rather than E-I estimation. In addition, these models used structural connectivity (SC) from Diffusion Tensor Imaging (DTI) as a proxy for synaptic efficiency, assumed the same local kinetic parameters for all regions and estimated only one global scaling coefficient for all long-range inter-regional connections, which cannot infer region-specific E-I imbalance in AD.

To overcome the aforementioned limitations of existing modeling studies, we applied a recently developed computational framework termed “Multiscale Neural Model Inversion (MNMI)” (Li et al., 2019; 2021) to the rs-fMRI data obtained from the Alzheimer’s Disease Neuroimaging Initiative (ADNI) database to identify region-specific E-I imbalance in AD. Compared with other major generative modeling frameworks such as Dynamic Causal Modeling (DCM; Friston et al., 2003, 2014; Li et al., 2011) and Biophysical Network Model (BNM; Honey et al., 2007, 2009; Deco and Jirsa, 2012; Deco et al., 2013a, b), the strengths of MNMI include using a biologically plausible neural mass model to describe network dynamics, estimating both intra-regional and inter-regional effective connectivity (EC), and constraining EC estimation with structural information. Specifically, MNMI estimates within-region (local) recurrent excitation and inhibition coupling weights as well as inter-regional connection strengths at single subject level based on rs-fMRI, thus enabling the inference of region-specific E-I balance. We focused our analysis on four functional networks (DMN, salience, executive control and limbic) due to their critical role and significant disruption in AD pathophysiology (Lustig et al., 2003; Greicius et al., 2004; Menon, 2011; Dhanjal and Wise, 2014; Badhwar et al., 2017; Schultz et al., 2017). Results indicated that MNMI is able to identify altered regional E-I balance in MCI and AD which deteriorates with disease progression and correlates with cognitive performance. This computational study offers mechanistic insights into the alteration of E-I balance during AD progression and the findings have the potential to contribute to the development of novel diagnostic techniques and treatment approaches by enabling the detection and modulation of E-I imbalance in AD.

## Results

To identify E-I imbalance in AD, we applied the MNMI model (Fig. 1) to a rs-fMRI dataset from ADNI consisting of 48 normal control (NC), 48 MCI and 48 AD subjects. At the heart of the MNMI framework is a neural mass model consisting of multiple brain regions each containing one excitatory and one inhibitory neural populations coupled with reciprocal connections. The excitatory neural populations are interconnected with long-range fibers whose baseline connection strengths are determined by structural connectivity (SC) from diffusion MRI. The neural activities are converted to simulated BOLD signals via a hemodynamic response function (HRF) and simulated FC is computed. MNMI then estimates both intra-regional and inter-regional connection strengths using genetic algorithm to minimize the difference between simulated and empirical FC (Fig. 1). We constructed a large network model with 46 regions of interest (ROIs) selected from the DMN, salience, executive control and limbic/subcortical networks (Table 1), and used the DTI data of 100 unrelated subjects from the Human Connectome Project (HCP) to calculate the baseline SC matrix. After model connection parameters (i.e., EC) are estimated for each individual subject, we derived regional E-I balance based on incoming excitatory and inhibitory connection strengths. We next performed statistical analysis to identify disrupted EC and E-I balance in MCI and AD. Lastly, we examined the association between E-I difference (ratio) and cognitive performance represented by the Mini-Mental State Examination (MMSE) score.

**Figure 1.**
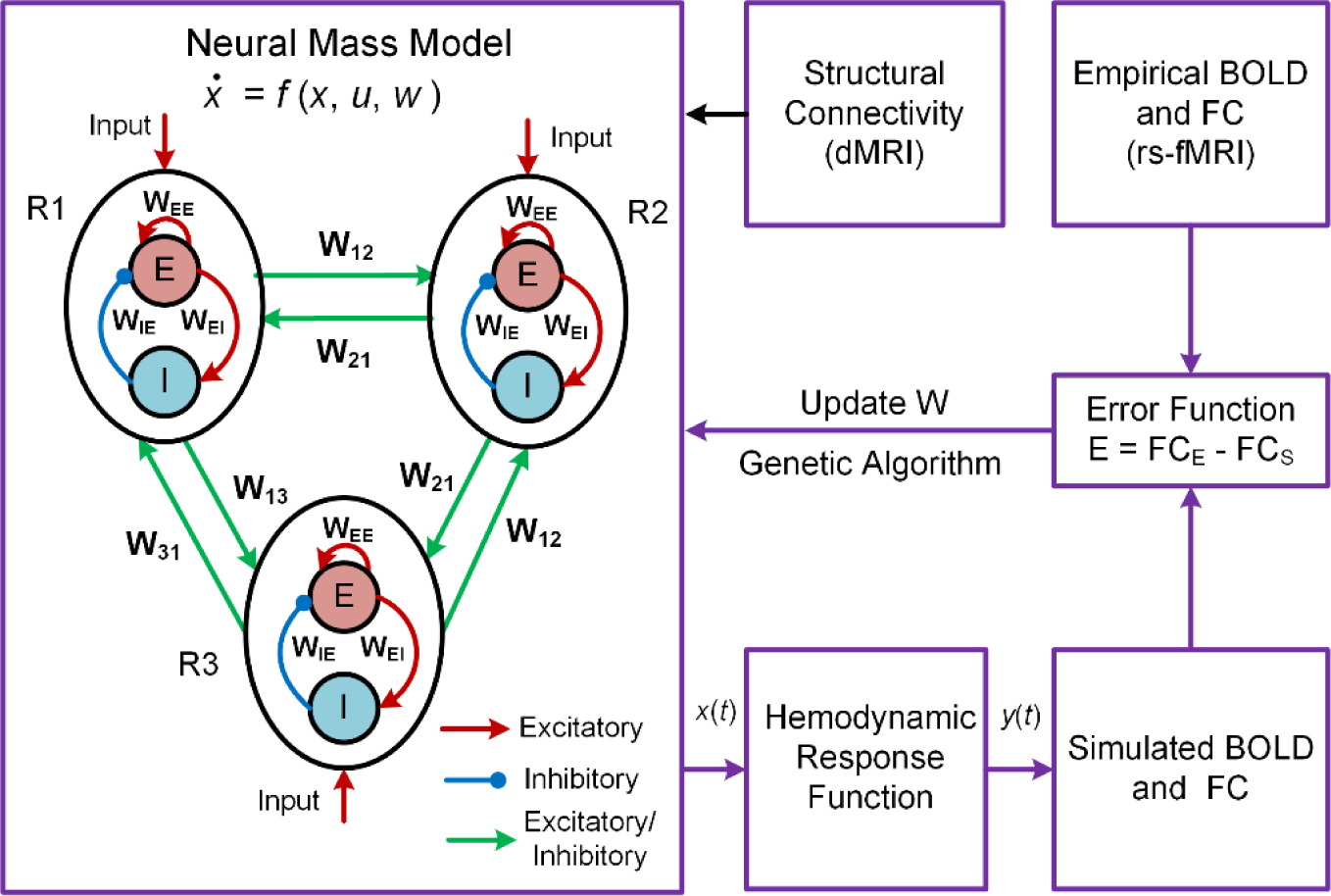
Overview of the MNMI framework. The neural activity (*x*) is described by a neural mass network model containing multiple brain regions (R1, R2, etc.). Each region consists of one excitatory (*E*) and one inhibitory (*I*) neural population with reciprocal connections. Inter-regional connection strength is based on SC from diffusion MRI. The neural activity (*x*) is converted to corresponding BOLD signals *(y*) via a hemodynamic response function. The model parameters are optimized to minimize the difference between simulated FC and empirical FC obtained from rs-fMRI.

**Table 1.**
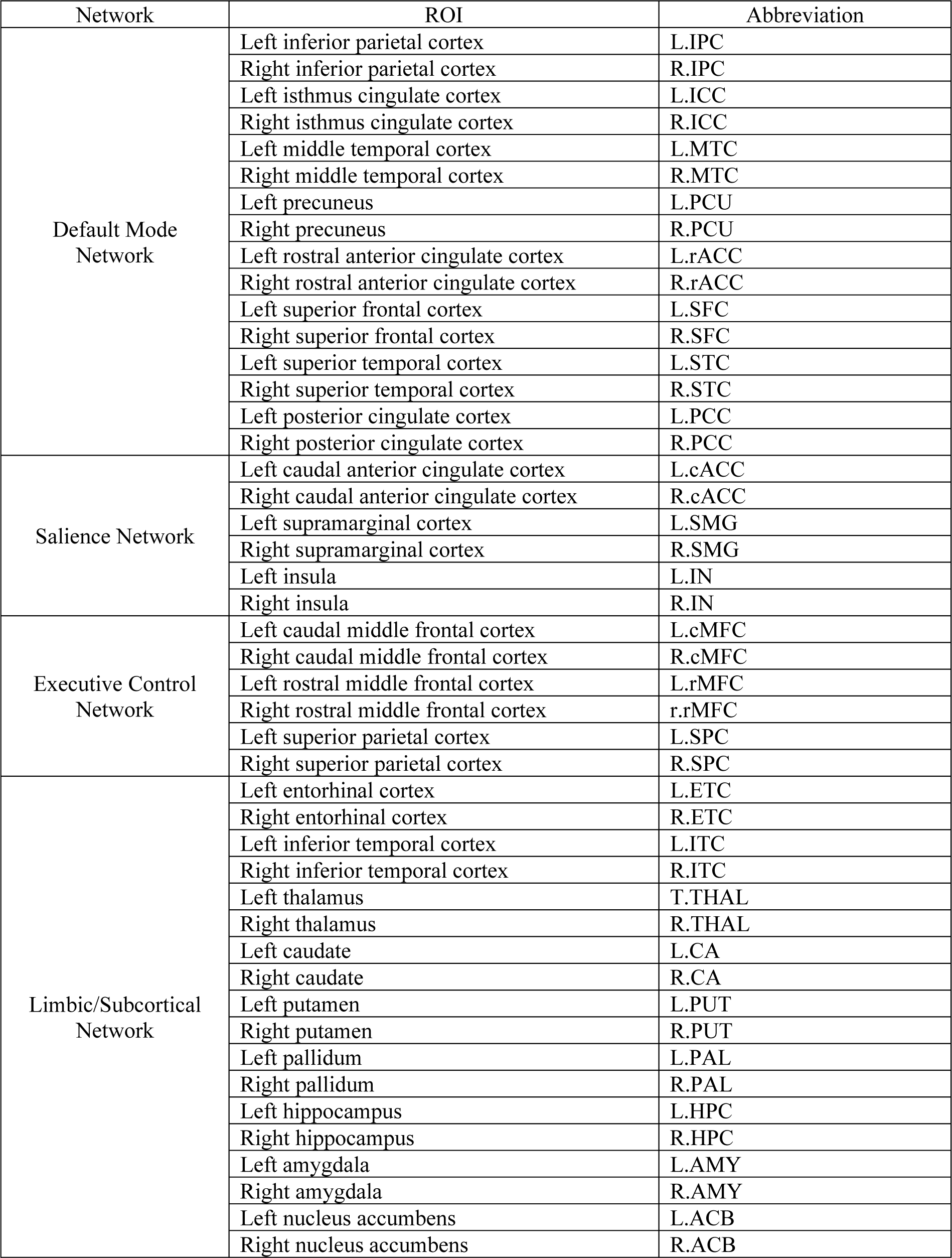
Regions of interest (ROIs) selected in network modeling.

| Network | ROI | Abbreviation |
| --- | --- | --- |
| Default Mode Network | Left inferior parietal cortex | L.IPC |
|  | Right inferior parietal cortex | R.IPC |
|  | Left isthmus cingulate cortex | L.ICC |
|  | Right isthmus cingulate cortex | R.ICC |
|  | Left middle temporal cortex | L.MTC |
|  | Right middle temporal cortex | R.MTC |
|  | Left precuneus | L.PCU |
|  | Right precuneus | R.PCU |
|  | Left rostral anterior cingulate cortex | L.rACC |
|  | Right rostral anterior cingulate cortex | R.rACC |
|  | Left superior frontal cortex | L.SFC |
|  | Right superior frontal cortex | R.SFC |
|  | Left superior temporal cortex | L.STC |
|  | Right superior temporal cortex | R.STC |
|  | Left posterior cingulate cortex | L.PCC |
|  | Right posterior cingulate cortex | R.PCC |
| Salience Network | Left caudal anterior cingulate cortex | L.cACC |
|  | Right caudal anterior cingulate cortex | R.cACC |
|  | Left supramarginal cortex | L.SMG |
|  | Right supramarginal cortex | R.SMG |
|  | Left insula | L.IN |
|  | Right insula | R.IN |
| Executive Control Network | Left caudal middle frontal cortex | L.cMFC |
|  | Right caudal middle frontal cortex | R.cMFC |
|  | Left rostral middle frontal cortex | L.rMFC |
|  | Right rostral middle frontal cortex | r.rMFC |
|  | Left superior parietal cortex | L.SPC |
|  | Right superior parietal cortex | R.SPC |
| Limbic/Subcortical Network | Left entorhinal cortex | L.ETC |
|  | Right entorhinal cortex | R.ETC |
|  | Left inferior temporal cortex | L.ITC |
|  | Right inferior temporal cortex | R.ITC |
|  | Left thalamus | T.THAL |
|  | Right thalamus | R.THAL |
|  | Left caudate | L.CA |
|  | Right caudate | R.CA |
|  | Left putamen | L.PUT |
|  | Right putamen | R.PUT |
|  | Left pallidum | L.PAL |
|  | Right pallidum | R.PAL |
|  | Left hippocampus | L.HPC |
|  | Right hippocampus | R.HPC |
|  | Left amygdala | L.AMY |
|  | Right amygdala | R.AMY |
|  | Left nucleus accumbens | L.ACB |
|  | Right nucleus accumbens | R.ACB |

### MNMI performance

The performance of MNMI is illustrated in Fig. 2. The average fitness value (i.e., Pearson’s correlation between simulated and empirical FC) was 0.6 ± 0.08 for NC, 0.61 ± 0.07 for MCI, and 0.62 ± 0.08 for AD, respectively. Both the simulated neural activity and simulated BOLD signals displayed rhythmic fluctuations (Fig. 2A, B). The oscillation frequency of the neural activity was about 7-10 Hz, consistent with α oscillations during relaxed wakefulness (Hughes and Crunelli, 2005). The frequency of the BOLD signals ranged between 0.01 and 0.05 Hz, in line with experimental observation (Tong et al., 2019). The empirical and simulated FC are displayed in Fig. 2C, D respectively, where the pattern of the simulated FC closely matched that of the empirical FC (correlation coefficient 0.68).

**Figure 2.**
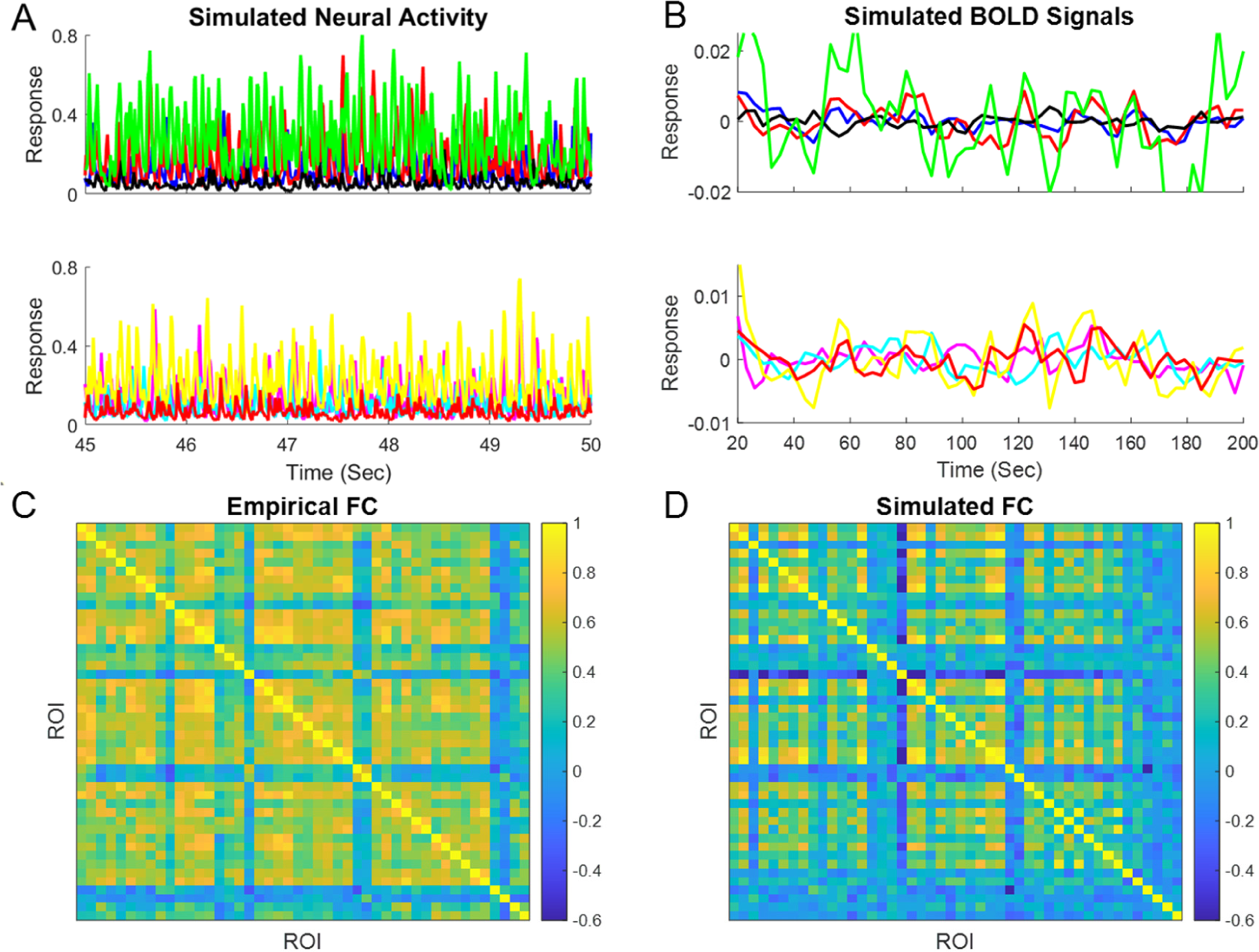
Performance of MNMI. **(A)** Sample activity of excitatory neural populations in eight randomly selected ROIs (four in top and four in bottom). **(B)** Sample BOLD signals in the same eight ROIs. **(C)** Empirical FC from a randomly selected subject. **(D)** Simulated FC from the same subject.

### Disrupted intra-regional E-I balance in MCI and AD

We first examined whether intra-regional (local) E-I balance was altered in MCI and AD. The recurrent excitation weights within 46 ROIs are shown in Fig. 3A1 for NC and MCI, Fig. 3A2 for NC and AD, and Fig. 3A3 for MCI and AD respectively. For the NC to MCI comparison of recurrent excitation, three regions (R.IPC, L.rACC and R.cACC) showed marginally significant decrease (*p* < 0.05, uncorrected), and three regions (R.ETC, L.PAL and R.ACB) showed marginally significant increase (*p* < 0.05, uncorrected) (Fig. 3A1). For NC versus AD, six regions displayed significant difference in recurrent excitation, including L.cMFC, R.PAL, L.HPC and L.AMY with decreased excitation, and L.PCC and R.ACB with increased excitation (Fig. 3A2). The significant excitation increase in R.ACB survived correction for multiple comparisons (*p* < 0.05, FDR corrected), and the R.ACB was the only region that showed significant and consistent excitation change across both MCI and AD. Regarding the comparison of MCI versus AD, only the region of R.AMY showed marginally significant decrease in AD (*p* < 0.05, uncorrected; Fig. 3A3). This suggests that as a prodromal stage of AD, MCI has similar recurrent excitation levels as AD, though AD shows more significant impairment in certain brain regions (e.g., R.ACB) when compared with NC.

**Figure 3.**
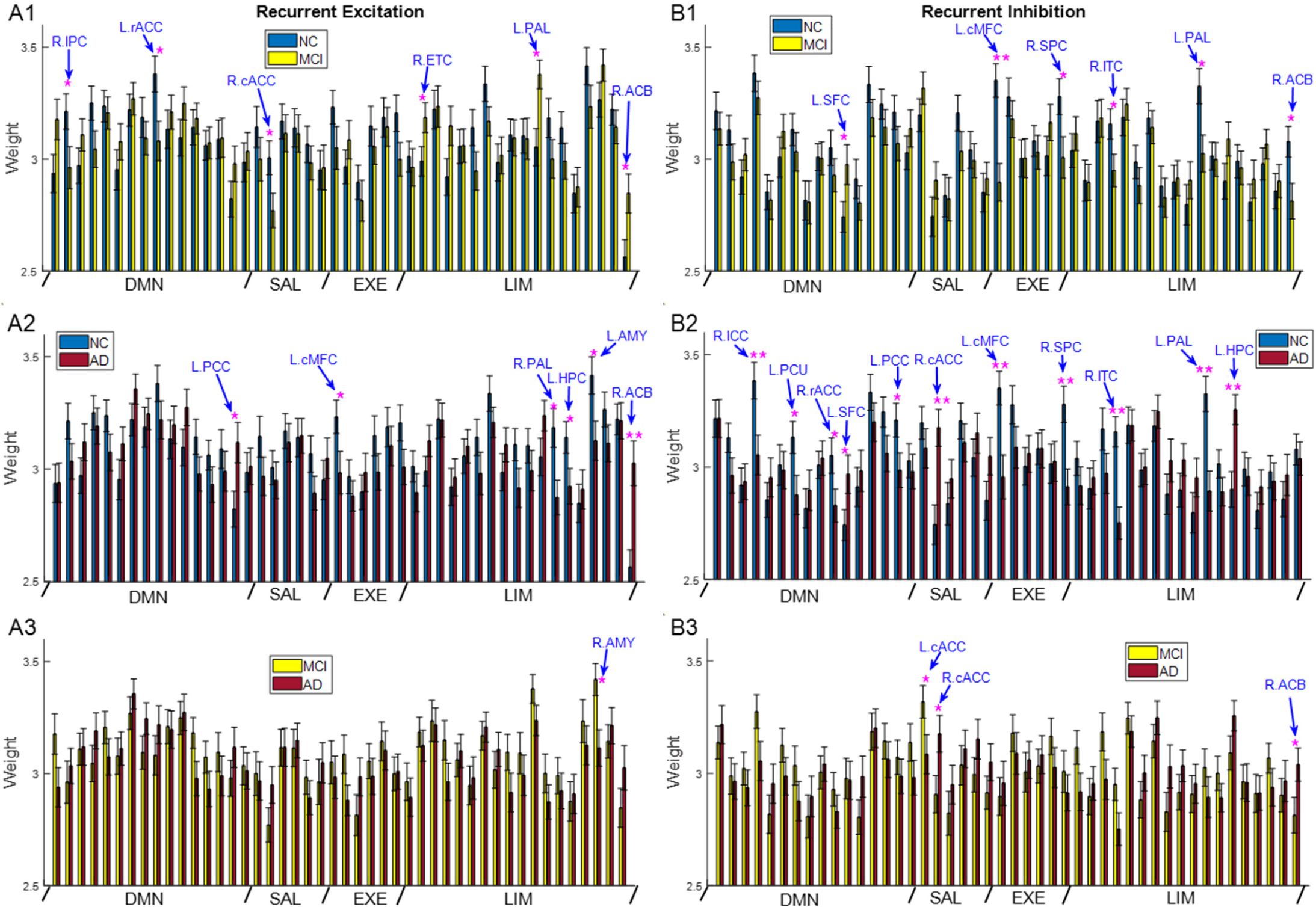
Impaired local recurrent excitation and inhibition in MCI and AD. Comparison of average recurrent excitation weight between NC and MCI (**A1**), NC and AD (**A2**), and MCI and AD (**A3**) for all 46 ROIs. Comparison of average recurrent inhibition weight between NC and MCI (**B1**), NC and AD (**B2**), and MCI and AD (**B3**) for all 46 ROIs. One star indicates uncorrected significance (*p* < 0.05) and double stars indicate corrected significance by FDR (*p* < 0.05). DMN: default mode network, SAL: salience network, EXE: executive control network, LIM: limbic network. Error bars indicate standard errors.

Comparison of the recurrent inhibition weights within 46 ROIs is shown in Fig. 3B1 for NC and MCI, Fig. 3B2 for NC and AD, and Fig. 3B3 for MCI and AD, respectively. Both MCI and AD showed significant difference in recurrent inhibition compared to NC. In MCI, the recurrent inhibition weight of five regions (L.cMFC, R.SPC, R.ITC, L.PAL and R.ACB) was significantly decreased (*p* < 0.05) while one region (L.SFC) showed significant increase (*p* < 0.05; Fig. 3B1). In particular, the change in L.cMFC survived multiple comparison correction (*p* < 0.05, FDR corrected). In AD, the change in recurrent inhibition was much more pronounced than in MCI (Fig. 3B2). Specifically, eleven ROIs exhibited a significant difference in AD compared with NC, and the change in seven ROIs passed multiple comparison correction. The recurrent inhibition of R.ICC, L.PCU, R.rACC, L.PCC, L.cMFC, R.SPC, R.ITC and L.PAL was significantly reduced, while that of L.SFC, R.cACC and L.HPC was significantly increased in AD. The regions that survived multiple comparison correction included R.ICC, R.cACC, L.cMFC, R.SPC, R.ITC, L.PAL and L.HPC (Fig. 3B2, marked by double pink stars). Notably, five ROIs showed consistent change in recurrent inhibition across both MCI and AD (compare Fig. 3B1 with Fig. 3B2), specifically L.SFC, L.cMFC, R.SPC, R.ITC and L.PAL. Importantly, the difference in R.SPC, R.ITC and L.PAL was only marginally significant in MCI, but was robust to correction for multiple comparisons in AD, suggesting greater disruption of inhibitory interactions in AD than in MCI. Despite greater impairments of recurrent inhibition in AD than MCI, the difference between MCI and AD was only marginally significant in three ROIs including L.cACC, R.cACC, and R.ACB (Fig. 3B3). Again, this suggests that the changes in recurrent excitation and inhibition become more subtle from MCI to AD, compared with the changes from NC to MCI or AD.

To visualize the alterations in recurrent excitation and inhibition better, we listed the significant changes in MCI and AD (from NC) in Table 2 where an up arrow indicates a significant increase while a down arrow indicates a significant decrease. Several observations can be made. <u>First</u>, more connections were significantly different in AD than MCI. This is to be expected as AD represents a more severe disease stage than MCI. <u>Second</u>, the strength of the majority of connections (69%) was decreased in MCI/AD compared with NC. This is consistent with the widespread decrease in FC during the progression of AD (Filippi and Agosta, 2011; Brier et al., 2014; Dennis and Thompson, 2014). <u>Third</u>, if a region exhibited impairments in both recurrent excitation and inhibition, their directions of change were opposite to each other thus strengthening E-I imbalance, except for the executive control network where recurrent excitation and recurrent inhibition changed in the same direction. This suggests the existence of compensatory mechanisms in the executive control network to maintain similar E-I balance in the presence of AD pathology due to the critical role of this network in cognitive function (Miller et al., 2001; Petrides, 2005; Koechlin and Summerfield, 2007). <u>Lastly</u>, recurrent inhibition is more significantly disrupted by MCI/AD than recurrent excitation, in agreement with the emerging viewpoint of interneuron dysfunction in network impairments (Li et al., 2016; Palop and Mucke, 2016; Xu et al., 2020). The consistent impairment of recurrent inhibition across MCI and AD also suggests that inhibitory connections are a more stable biomarker of AD than excitatory connections.

**Table 2.** Alterations in recurrent excitation and inhibition strength in MCI and AD (compared with NC). Up arrows indicate increase and down arrows indicate decrease. One star indicates uncorrected significance and double stars indicate corrected significance by FDR.

| Network | ROI | MCI |  | AD |  |
| --- | --- | --- | --- | --- | --- |
|  |  | Excitation | Inhibition | Excitation | Inhibition |
| DMN | R.IPC | * ↓ |  |  |  |
|  | R.ICC |  |  |  | ** ↓ |
|  | L.PCU |  |  |  | * ↓ |
|  | L.rACC | * ↓ |  |  |  |
|  | R.rACC |  |  |  | * ↓ |
|  | L.SFC |  | * ↑ |  | * ↑ |
|  | L.PCC |  |  | * ↑ | * ↓ |
| SAL | R.cACC | * ↓ |  |  | ** ↑ |
| EXE | L.cMFC |  | ** ↓ | * ↓ | ** ↓ |
|  | R.SPC |  | * ↓ |  | ** ↓ |
| LIM | R.ETC | * ↑ |  |  |  |
|  | R.ITC |  | * ↓ |  | ** ↓ |
|  | L.PAL | * ↑ | * ↓ |  | ** ↓ |
|  | R.PAL |  |  | * ↓ |  |
|  | L.HPC |  |  | * ↓ | ** ↑ |
|  | L.AMY |  |  | * ↓ |  |
|  | R.ACB | * ↑ | * ↓ | ** ↑ |  |

The alteration in recurrent excitation and inhibition strengths resulted in intra-regional E-I imbalance in MCI and AD as shown in Fig. 4. The intra-regional (local) E-I balance was quantified as the E-I difference (i.e., recurrent excitation strength – recurrent inhibition strength; similar results were obtained for E/I ratio), which measures the level of net excitation. In MCI, three regions showed a significant decrease in intra-regional E-I difference without passing multiple comparison correction, including L.rACC, R.cACC and L.HPC (Fig. 4A). Three other regions in the limbic network displayed significant increase in intra-regional E-I difference, including R.ITC, L.PAL and R.ACB, among which the elevation within L.PAL and R.ACB passed multiple comparison correction. In AD, five regions showed consistent E-I impairments as MCI, including R.cACC, R.ITC. L.PAL, L.HPC and R.ACB (Fig. 4B). Notably, the E-I alteration in R.cACC and L.HPC became more robustly significant in AD than MCI, surviving multiple comparison correction (the corrected significance in L.PAL and R.ACB maintained as MCI). In addition to the five common ROIs, intra-regional E-I balance in L.PCU, L.PCC, L.PUT and L.AMY were also impaired in AD, with a significant increase in L.PCU and L.PCC, and a significant decrease in L.PUT and L.AMY (*p* < 0.05). The increase in L.PCC was able to pass correction for multiple comparisons (*p* < 0.05, FDR corrected). In contrast to the marked E-I balance changes from NC to MCI/AD, the changes from MCI to AD were much less pronounced with only three ROIs showing uncorrected significance (R.ICC, R.SFC, and L.PUT; Fig. 4C). Specifically, the E-I difference of R.SFC and L.PUT was significantly decreased (*p* < 0.05, uncorrected), while that of R.ICC was significantly increased (*p* < 0.05, uncorrected). Overall, intra-regional E-I imbalance in MCI and AD was highly consistent and concentrated on the limbic network (Fig. 4A, B). Also, in MCI/AD, about half of the brain regions showed increased intra-regional E-I difference while the other half exhibited reduced E-I difference when compared with NC. To visualize the progressive changes in E-I balance from NC to MCI and to AD, we fit a linear model to the local E-I difference of all NC, MCI and AD subjects and found the model significance of six brain regions passed correction for multiple comparison (*p* < 0.05, FDR corrected) including L.PCC, R.cACC, R.ITC, L.PAL, L.HPC, and R.ACB. Specifically, the E-I difference of L.PCC, R.ITC, L.PAL and R.ACB was progressively increased from NC to MCI and to AD, while the E-I difference of R.cACC and L.HPC was progressively decreased from NC to MCI and to AD. Notably, the brain regions that survived multiple comparison correction for the linear model were consistent with those regions showing common significant changes across MCI and AD (Fig. 4A, B), except for L.PCC which displayed significant change in AD only.

**Figure 4.**
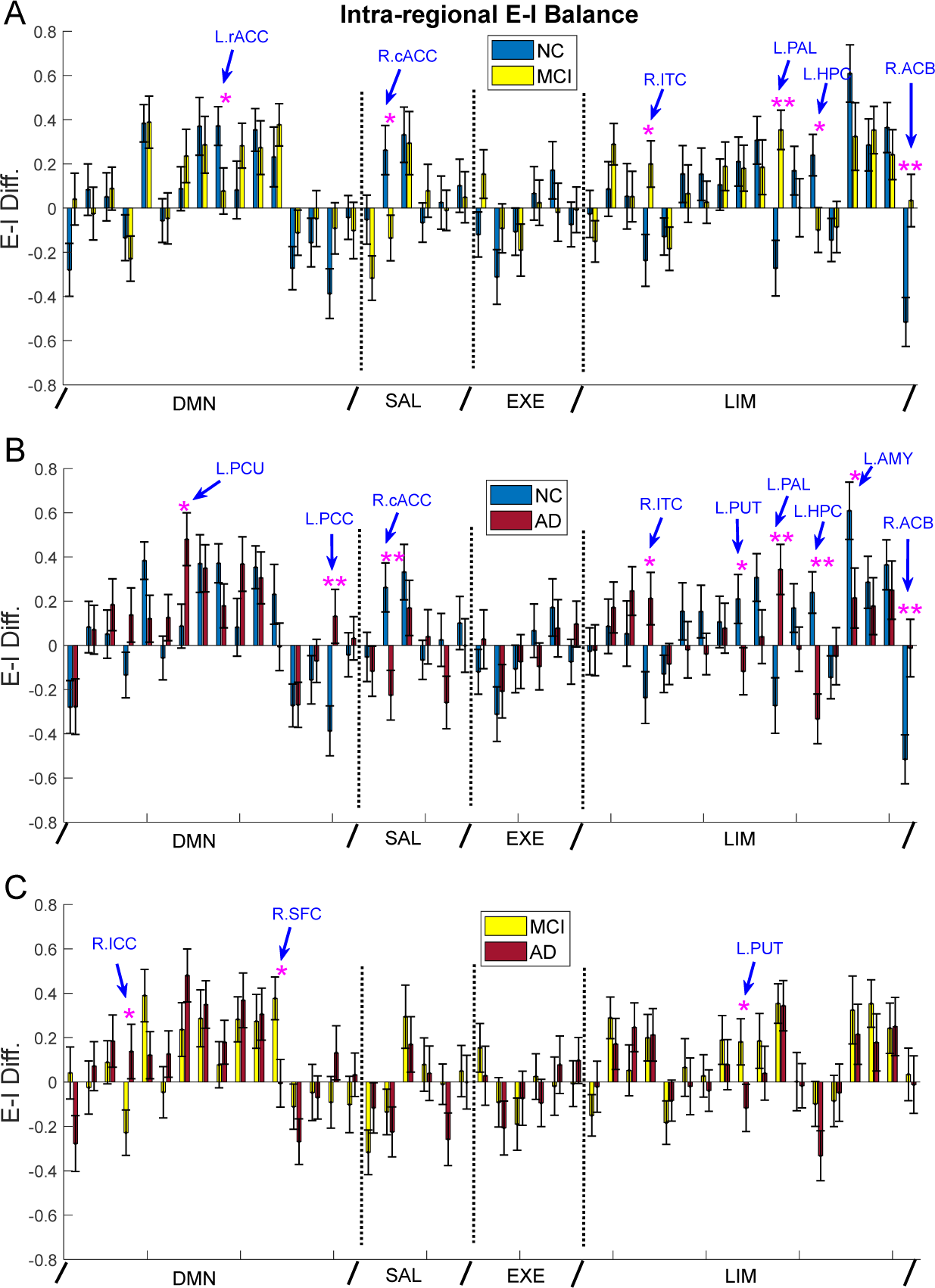
Disrupted intra-regional E-I balance in MCI and AD. Comparison of local E-I difference (recurrent excitation – recurrent inhibition) between NC and MCI (**A**), NC and AD (**B)**, and MCI and AD (**C**) for all 46 ROIs. One star indicates uncorrected significance and double stars indicate corrected significance by FDR (*p* < 0.05).

We next examined network-averaged recurrent excitation and inhibition changes in MCI and AD. There was no significant difference between NC and MCI for recurrent excitation (Fig. 6A) while the executive control network showed decreased recurrent inhibition in MCI compared with NC (*p* < 0.05, uncorrected; Fig. 6B). By comparison, significant reduction in both recurrent excitation and inhibition was observed in the executive control network in AD and the change in recurrent inhibition survived multiple comparison correction (Fig. 6A, B). Moreover, the DMN exhibited a reduction in recurrent inhibition while the salience network showed an increase in recurrent inhibition (*p* < 0.05, uncorrected; Fig. 6B). Thus, on the network level, impairments in recurrent excitation and inhibition became more notable in AD than in MCI and the executive control network showed the most significant and consistent alterations. The decrease in both recurrent excitation and inhibition may compensate for the loss of each other, thus maintaining a relatively stable E-I balance in the executive control network. Lastly, no significant difference was observed between MCI and AD, indicating a similar impairment level at the network level.

**Figure 5.**
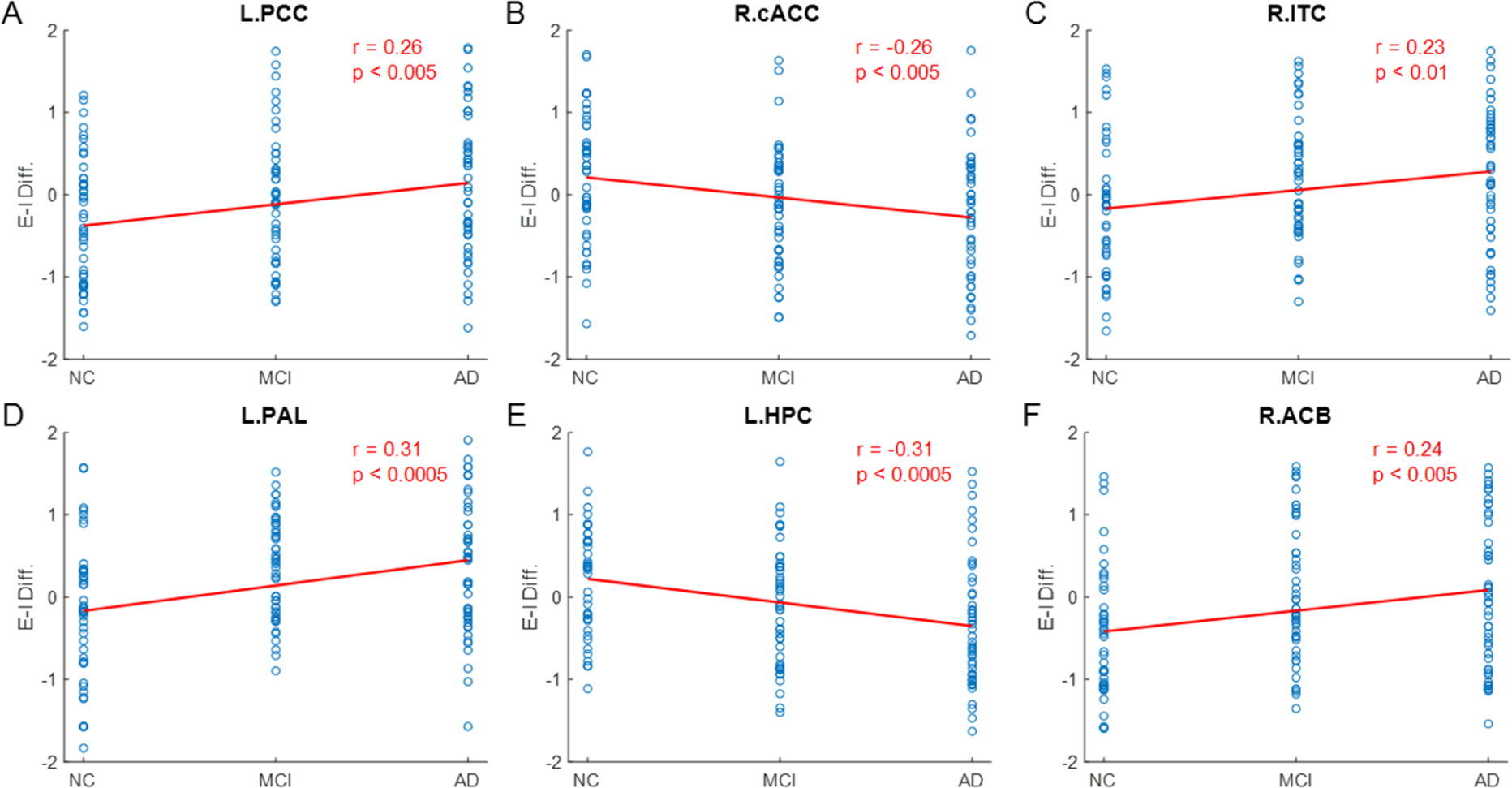
Progressive changes in intra-regional E-I balance from NC to MCI and to AD. Local E-I difference of NC, MCI and AD subjects is fit by a linear model for (**A**) L.PCC, (**B**), R.cACC, (**C**) R.ITC, (**D**) L.PAL, (**E**) L.HPC, and (**F**) R.ACB. The significance of the linear fit for all six ROIs passes multiple comparison correction by FDR (*p* < 0.05).

**Figure 6.**
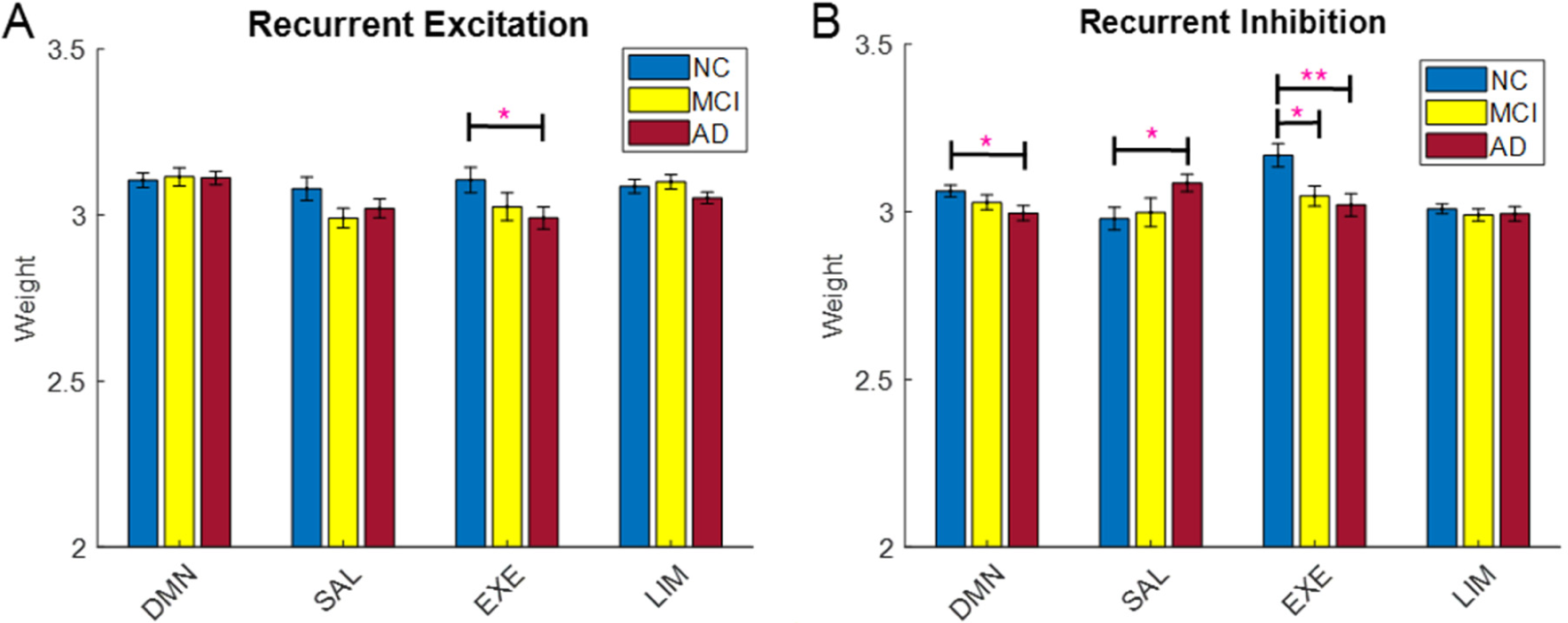
Network-averaged recurrent excitation and inhibition weight. **(A)** Comparison of average recurrent excitation weight within four functional networks among NC, MCI and AD. **(B)** Comparison of average recurrent inhibition weight within four functional networks among NC, MCI and AD. One star indicates uncorrected significance and double stars indicate corrected significance by FDR (*p* < 0.05).

### Disrupted inter-regional E-I balance in MCI and AD

In addition to intra-regional recurrent connections, inter-regional excitatory and inhibitory connections were also disrupted in MCI and AD. The color-coded average inter-regional EC matrices for NC, MCI and AD are shown in Fig. 7A, B, and C, respectively (the white area indicates removed weak connections). We observed that the EC patterns were similar for NC, MCI and AD where there were more excitatory (positive) connections than inhibitory (negative) connections. The significant EC connections in MCI (compared to NC) are indicated by the blue edges in Fig. 7D (*p* < 0.05, uncorrected), where they were broadly distributed among the four networks. The significant EC connections in AD (compared to NC) are shown in Fig. 7E where the blue edges indicate uncorrected significant connections and the red edges denote significant connections corrected by Network-based Statistics (NBS; Zalesky et al., 2010). Compared with MCI, the significant connections in AD concentrated more within and between the DMN and limbic networks. Of note, the corrected significant connections (red edges) involved mostly the executive control and limbic networks. Multiple significant connections also existed between MCI and AD comparison (*p* < 0.05, uncorrected; Fig. 7F), which mostly involved the DMN and executive control network. It should be noted that the overlap of significant connections among the three-way comparison (NC-MCI, NC-AD and MCI-AD) is low. To visualize the EC changes better, we compared the significant inter-regional EC between NC and MCI in Fig. 8A, between NC and AD in Fig. 8B, and between MCI and AD in Fig. 8C. As indicated by the EC difference in the bottom panels, most of the connections had less excitatory influence (or more inhibitory influence) in MCI and AD than NC (Fig. 8A, B), indicating less excitatory communication between regions in MCI and AD. The corrected significant connections in AD included R.SPC→R.PUT, R.CA→R.THAL, R.SFC→R.PAL, R.PAL→R.PUT, R.rMFC◊R.PUT, R.CA→R.rMFC, and R.PUT→R.PAL. Interestingly, most of the significant connections had increased EC in AD compared with MCI (Fig. 8C). This suggests that progression of AD may involve different sets of inter-regional connections that are differentially disrupted.

**Figure 7.**
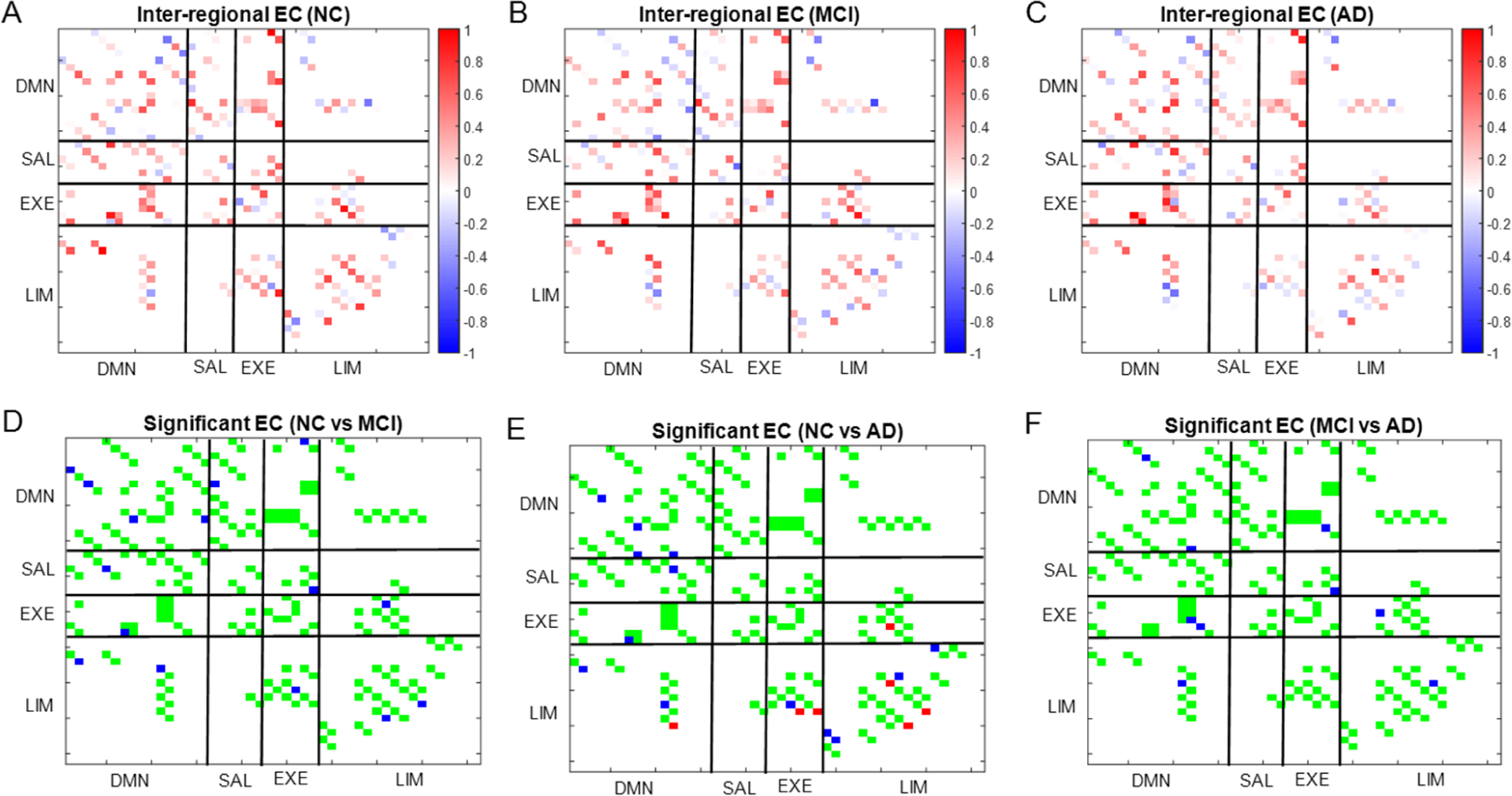
Altered inter-regional EC in MCI and AD. Average inter-regional EC matrix for NC (**A**), MCI (**B**), and AD (**C**)**. (D)** Significant EC connections in MCI when compared with NC. **(E)** Significant EC connections in AD when compared with NC. **(F)** Significant EC connections in AD when compared with MCI. For **(D-F)**, green edges indicate insignificant connections, blue edges indicate uncorrected significant connections (*p* < 0.05), and red edges indicate significant connections corrected by the Network-based Statistics (NBS; *p* < 0.05).

**Figure 8.**
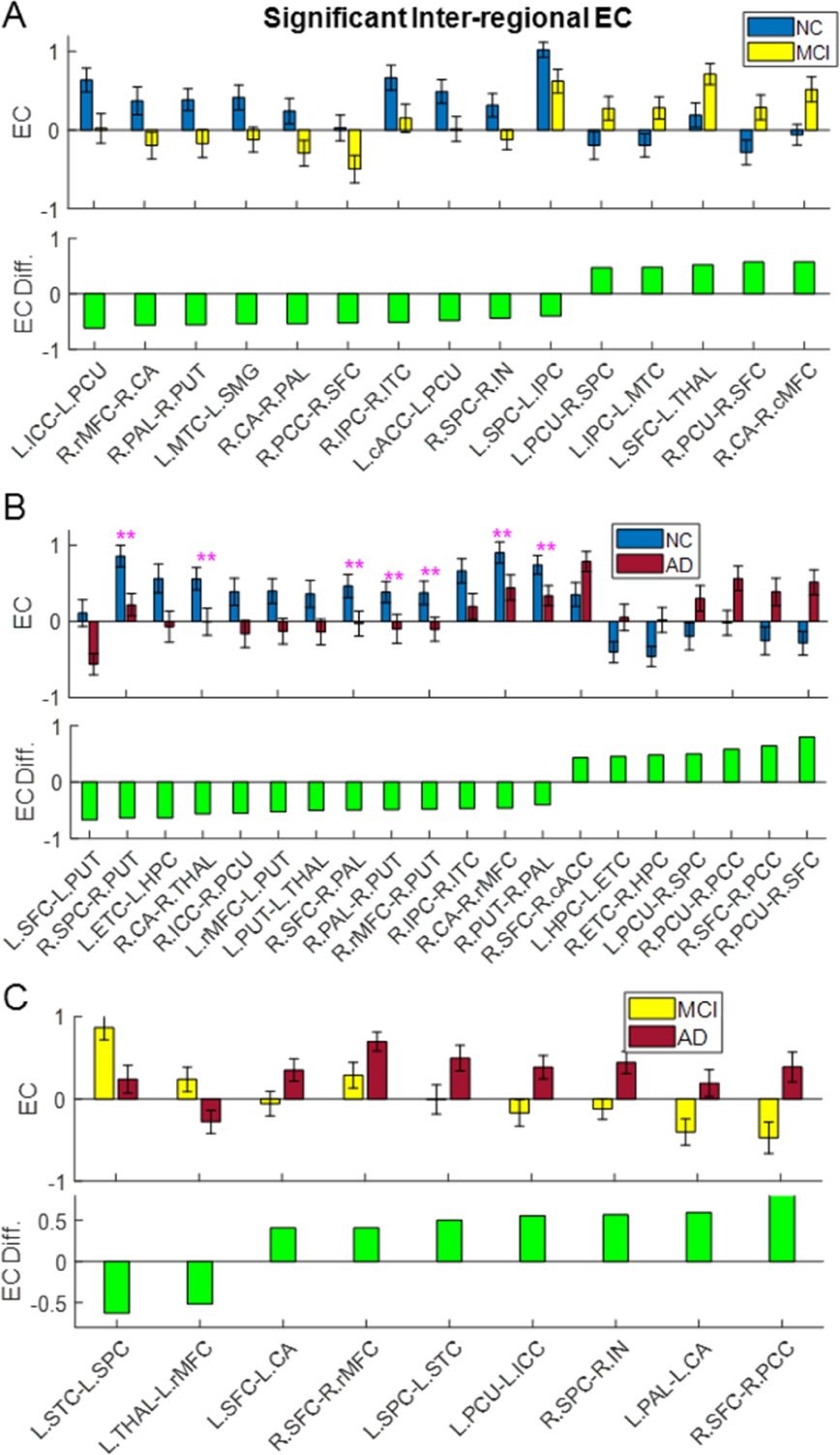
Significant inter-regional connections for group pairwise comparison. Significant inter-regional EC for NC and MCI comparison (**A**), NC and AD comparison (**B**) and MCI and AD comparison (**C**). The top panel plots the average inter-regional EC while the bottom panel plots the EC difference between the two compared groups. Double stars indicate corrected significance by NBS *(p* < 0.05).

To evaluate the inter-regional E-I balance change in MCI and AD, we computed the inter-regional E-I difference (i.e., difference between the sum of all positive incoming EC and the absolute sum of all negative incoming EC) to a particular ROI (Fig. 9). The E-I change from NC to MCI is shown in Fig. 9A where six regions showed impaired inter-regional E-I balance (*p* < 0.05, uncorrected). A majority of the six ROIs showed decreased excitation, including L.PCU, R.IN, R.ITC, R.PUT and R.PAL, and only one ROI (R.cMFC) exhibited increased excitation. In AD, three common regions showed reduced net excitation as MCI, including R.ITC, R.PUT and R.PAL, all belonging to the limbic network (Fig. 9B). In particular, the significant changes in R.PUT and R.PAL were able to survive correction for multiple comparison by FDR, again indicating more severe E-I disruption in AD than MCI. Moreover, the E-I difference of L.HPC was significantly reduced (*p* < 0.05), while that of R.PCC was significantly elevated in AD (*p* < 0.05), both without passing multiple comparison correction. Comparison of the E-I difference between MCI and AD indicated that four brain regions had significant E-I balance changes (*p* < 0.05, uncorrected, Fig. 9C). Specifically, the E-I difference of L.IN, L.rMFC, and L.THAL was significantly reduced while that of R.PCC was significantly increased. Of note, the E-I difference of R.PCC was also significantly elevated in AD when compared with NC (Fig. 9B). We then fit a linear model to the E-I difference and found that the net excitation of R.PUT and R.PAL, the two regions with corrected significance in AD (Fig. 9B), reduced significantly from NC to MCI and to AD (*p* < 0.05, FDR corrected; Fig. 10). Overall, inter-regional E-I difference shows a decreasing trend over the course of AD progression, consistent with the majority of reduced inter-regional EC (Fig. 8).

**Figure 9.**
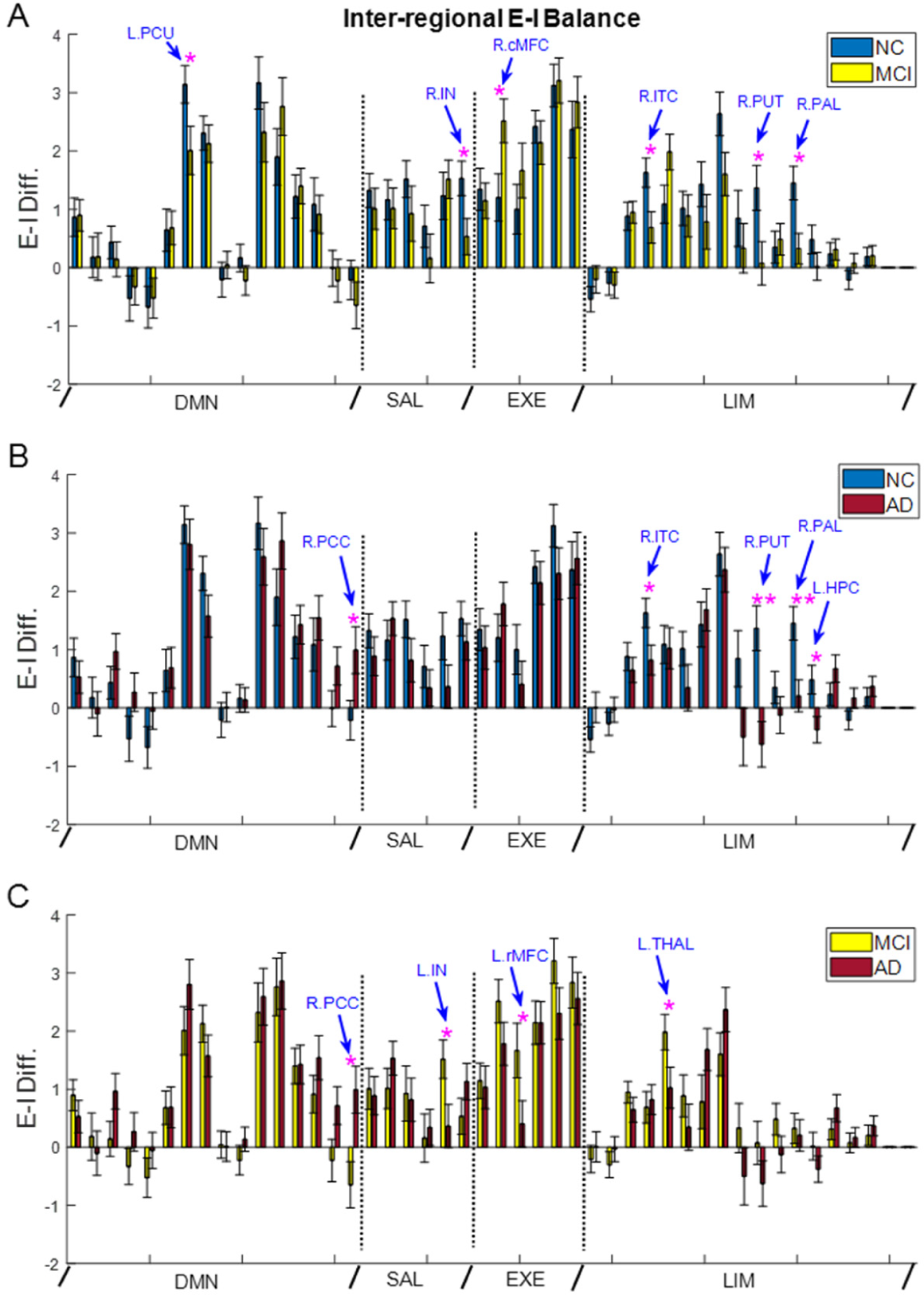
Disrupted inter-regional E-I balance in MCI and AD. Comparison of inter-regional E-I difference between NC and MCI (**A**), NC and AD (**B)**, and MCI and AD (**C**). One star indicates uncorrected significance and double stars indicate corrected significance by FDR *(p* < 0.05).

**Figure 10.**
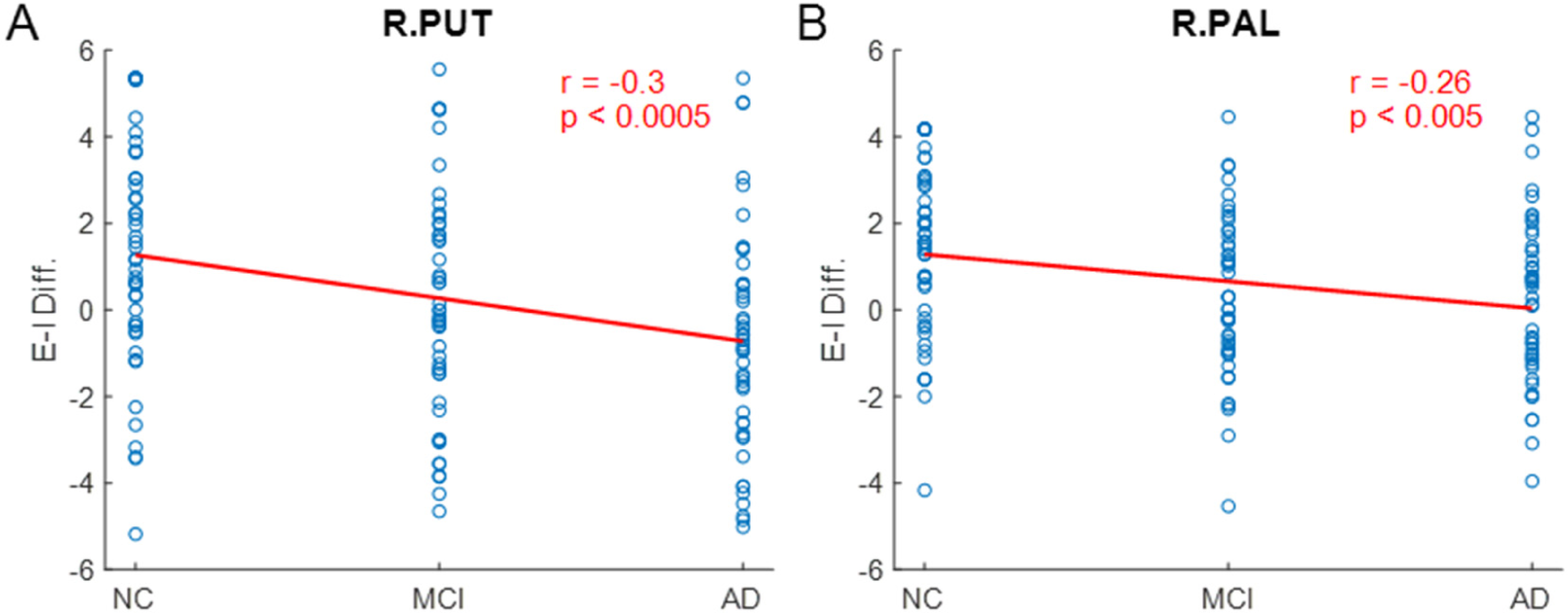
Progressive changes in inter-regional E-I balance from NC to MCI and to AD. Inter-regional E-I difference of NC, MCI and AD subjects is fit by a linear model for (**A**) R.PUT, and (**B**), R.PAL. The significance of the linear fit for both ROIs passes multiple comparison correction by FDR (*p* < 0.05).

The average inter-network EC (summation of all excitatory and inhibitory inter-regional EC between networks) is shown in Fig. 11. We observed that the EC from the executive control network to the salience network was significantly decreased in MCI when compared to NC (*p* < 0.05, uncorrected; Fig. 11A, B). In AD, the EC from the executive control network to the limbic network and the EC from the DMN to the limbic network were significantly reduced compared to NC with the latter passing multiple comparison correction (*p* < 0.05, FDR corrected) (Fig. 11A, C), suggesting cortical-limbic decoupling. Moreover, the EC from the limbic network to the executive network was significantly decreased from MCI to AD (*p* < 0.05, uncorrected; Fig. 11B, C). Overall, the excitatory interactions between networks are substantially decreased in MCI and AD when compared with NC, consistent with the predominant reduced inter-regional EC during AD progression (Fig. 8).

**Figure 11.**
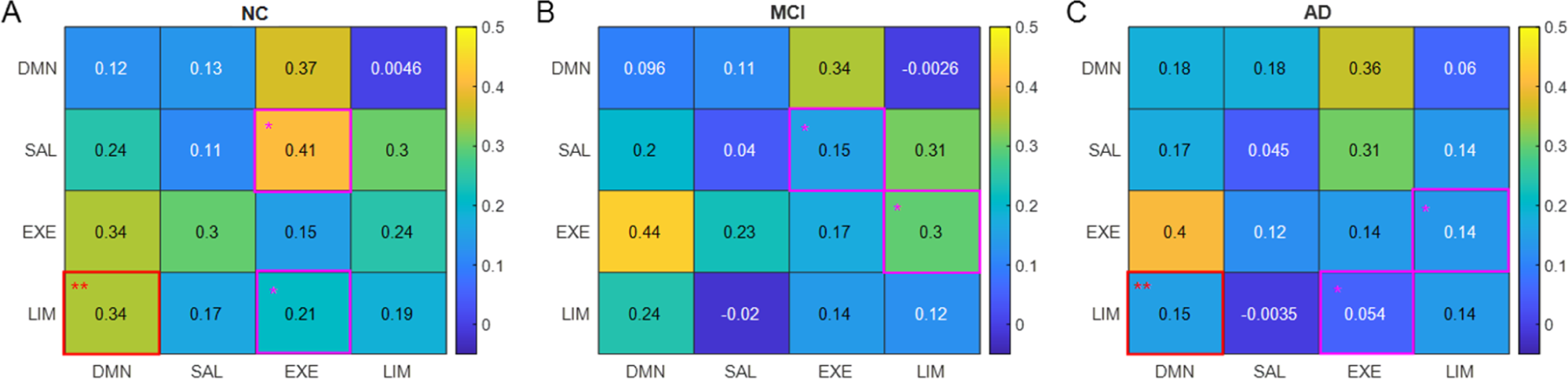
Altered inter-network connection strength in MCI and AD. **(A)** Average inter-network EC in NC. **(B)** Average inter-network EC in MCI. **(C)** Average inter-network EC in AD. Pink boxes (with one star) indicate uncorrected significance (*p* < 0.05) and red boxes (with double stars) indicate corrected significance (*p* < 0.05, corrected by FDR) for the corresponding connections.

### Disrupted overall E-I balance in MCI and AD

The above analysis indicates that both intra-regional and inter-regional E-I balance are impaired in MCI and AD. As the overall neural excitability depends on both intra-regional and inter-regional input drive, we computed the overall E-I balance as the difference between net excitation (recurrent excitation + all incoming excitatory inter-regional EC) and net inhibition (recurrent inhibition + all incoming inhibitory inter-regional EC) for all ROIs. We found that the overall E-I balance was altered in a number of regions in MCI and AD and most of the regions were located in the limbic network (Fig. 12). Specifically, the E-I difference of R.IN, R.PUT, R.PAL and L.HPC was significantly decreased (*p* < 0.05), while that of R.cMFC and R.ACB was significantly increased in MCI (*p* < 0.05); the change in R.PAL and R.ACB survived multiple comparison correction (Fig. 12A). In AD, the changes in overall E-I balance remained consistent for R.PUT, R.PAL, L.HPC and R.ACB, all four regions from the limbic network (compare Fig. 12B with 12A). In addition to R.PAL and R.ACB, the E-I balance changes in R.PUT and L.HPC were able to pass multiple comparison correction in AD. Besides, the overall E-I difference of L.PUT was significantly reduced, while that of L.PCC and R.PCC was significantly increased in AD (*p* < 0.05, Fig. 12B), all without surviving multiple comparison correction. Comparison of the overall E-I balance between MCI and AD revealed that four regions exhibited significant difference (*p* < 0.05, uncorrected), including L.ICC, L.PCC, R.PCC and L.IN (Fig. 12C). Specifically, the E-I difference of L.ICC, L.PCC and R.PCC were significantly increased, while that of L.IN was significantly reduced from MCI to AD. Of note, the E-I differences in L.PCC and R.PCC were also significantly increased from NC to AD (*p* < 0.05, uncorrected, Fig. 12B), but not from NC to MCI. This suggests that E-I disruption in PCC may be specific to the disease progression from MCI to AD and an elevation in excitability in the cingulate regions may signal the transition from MCI to AD. In contrast, significant E-I alterations in the limbic network (R.PUT, R.PAL, L.HPC and R.ACB) were observed in both NC-MCI and NC-AD comparison (Fig. 12A, B), but not in MCI-AD comparison (Fig. 12C). This indicates that E-I impairment in the limbic network was specific to NC to MCI progression and could be served as early biomarker for AD. Consistent with the common E-I alterations across MCI and AD, linear model analysis revealed the same four brain regions (out of 46 ROIs) in the limbic network (i.e., R.PUT, R.PAL, L.HPC and R.ACB) that exhibited significant progressive E-I changes from NC to MCI/AD (*p* < 0.05, FDR corrected; Fig. 13). Specially, the overall E-I difference in R.PUT, R.PAL and L.HPC was progressively reduced (Fig. 13A-C), while that of R.ACB was progressively increased from NC to MCI and to AD (Fig. 13D). Lastly, we examined the change of spontaneous input during AD progression. There was no difference in spontaneous input between NC and MCI, while the spontaneous input was significantly decreased in AD, when compared with NC or MCI (*p* < 0.05, FDR corrected; Fig. 14). This suggests that the overall excitatory drive to the network is reduced in the AD phase, consistent with overall reduction in E-I difference.

**Figure 12.**
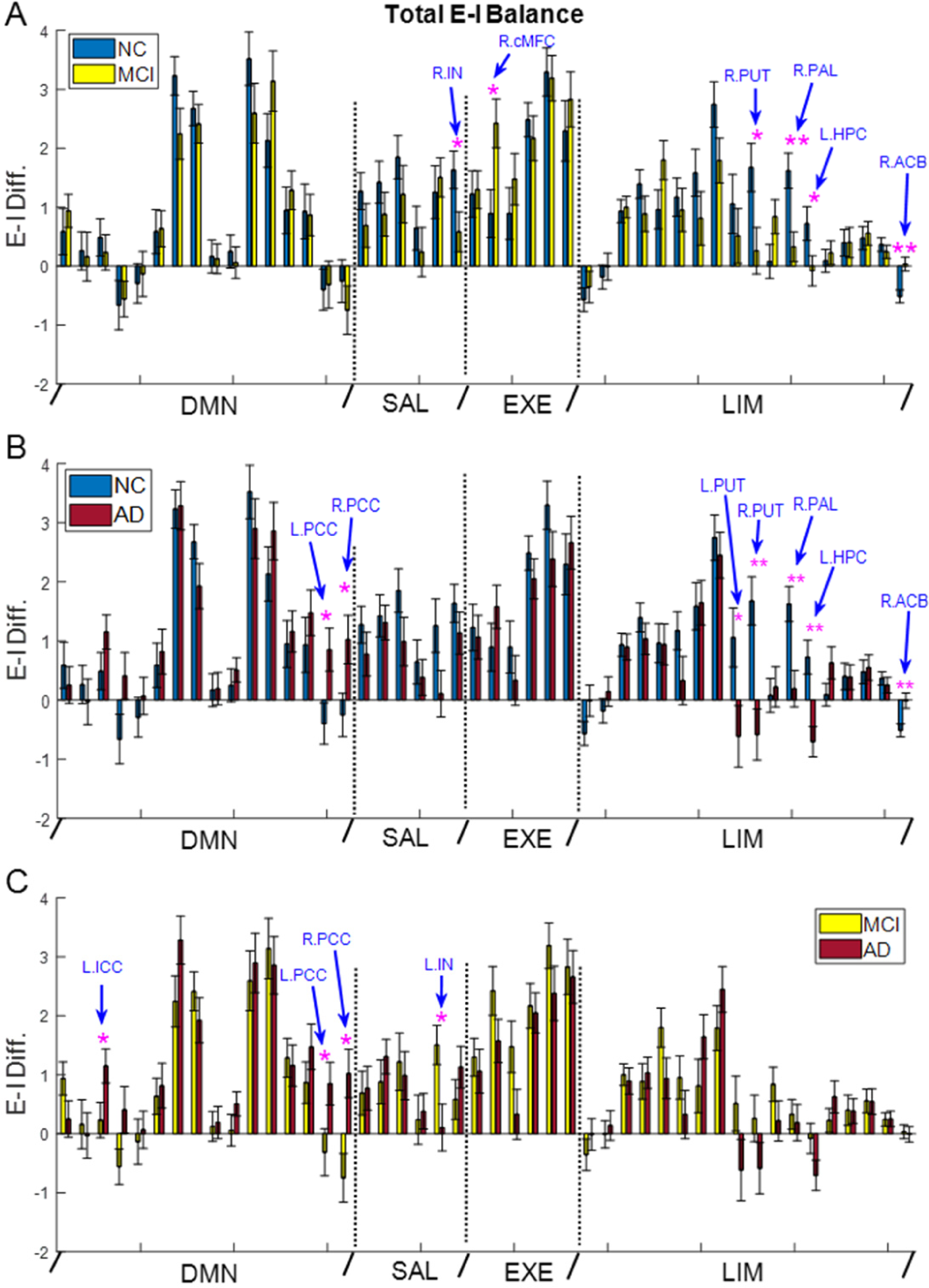
Disrupted overall E-I balance in MCI and AD. Comparison of total E-I difference between NC and MCI (**A**), NC and AD (**B**), and MCI and AD (**C**). One star indicates uncorrected significance and double stars indicate corrected significance by FDR *(p* < 0.05).

**Figure 13.**
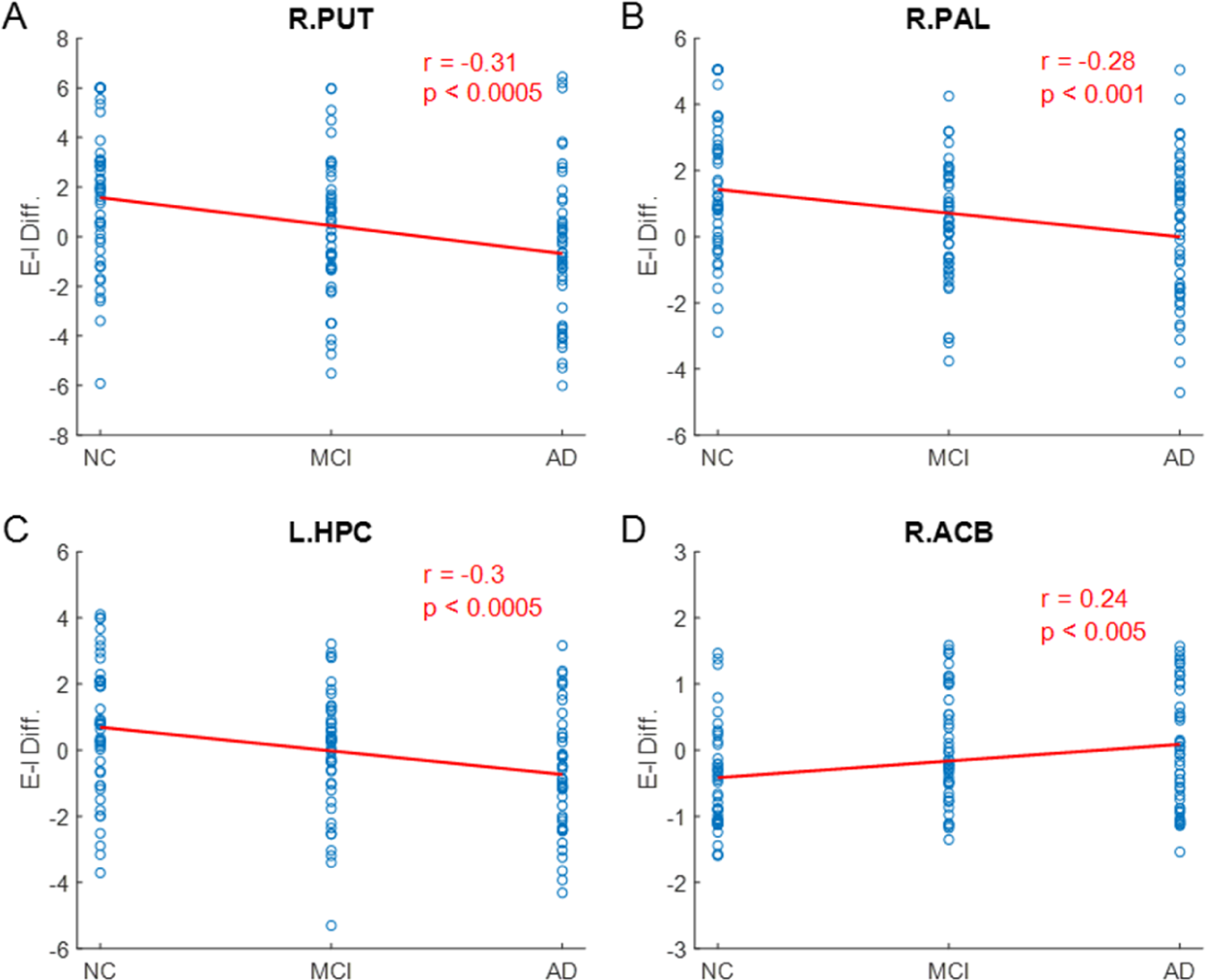
Progressive changes in overall E-I balance from NC to MCI and to AD. Total E-I difference of NC, MCI and AD subjects is fit by a linear model for (**A**) R.PUT, (**B**), R.PAL, (**C**) L.HPC and (**D**) R.ACB. The significance of the linear fit for all four ROIs passes multiple comparison correction by FDR (*p* < 0.05).

**Figure 14.**
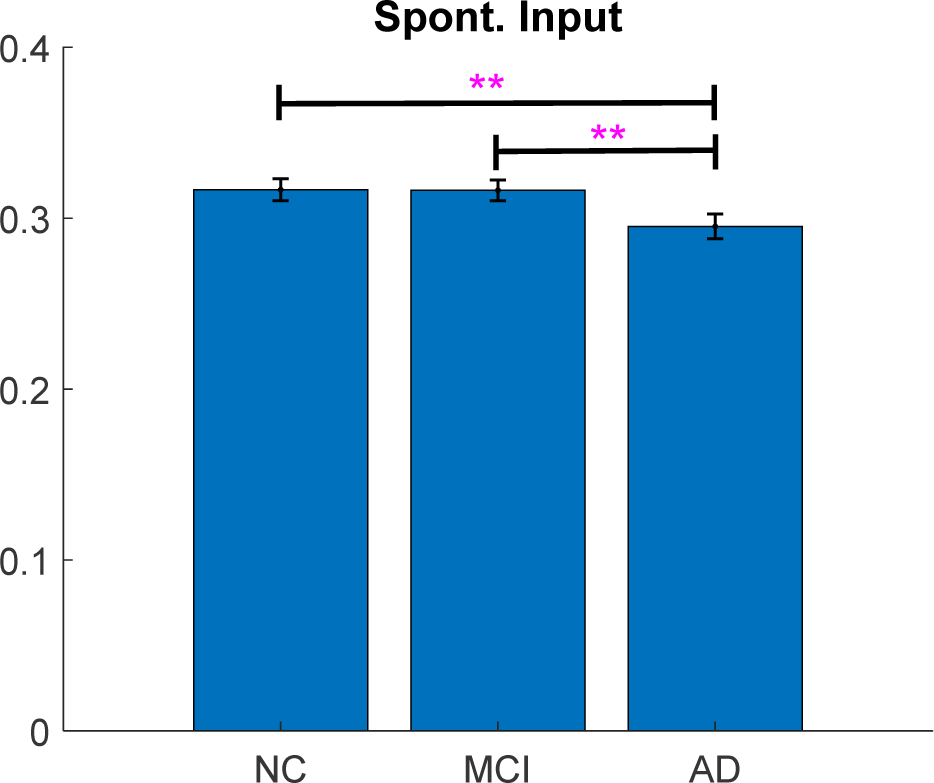
Comparison of estimated spontaneous inputs among NC, MCI and AD. Double stars indicate corrected significance by FDR *(p* < 0.05).

### Association between regional E-I balance and cognitive performance

To evaluate the behavioral relevance of E-I imbalance in AD progression, we examined the association between regional E-I balance and MMSE score. We performed the correlation analysis for intra-regional, inter-regional and total E-I balance respectively. The intra-regional and total E-I balance were evaluated as both the E-I difference (difference between E and I) and E/I ratio, while inter-regional E-I balance is assessed as the E-I difference only (see Methods). We computed the Pearson’s correlation between regional E-I balance and MMSE score for all 46 ROIs and the correlation that passed FDR correction (*p* < 0.05) is reported in Fig. 15. For intra-regional E-I balance, we observed that the E-I difference in L.HPC was positively correlated with the MMSE score (Fig. 15A), indicating that lower excitation in L.HPC was associated with lower MMSE score in MCI/AD. When intra-regional E-I balance was evaluated as E/I ratio, five regions showed significant correlation with MMSE score, including L.PCU, L.PCC, L.PAL, R.cACC, and L.HPC (Fig. 15B-F). Specifically, the E/I ratio in L.PCU, L.PCC and L.PAL was negatively correlated with MMSE score, while that in R.cACC and L.HPC was positively correlated with MMSE score. Thus, the higher intra-regional E/I ratio in L.PCU, L.PCC and L.PAL was associated with lower MMSE score in MCI/AD, which became oppositive for R.cACC and L.HPC. For total E-I balance, we found that the E-I difference in L.PUT, R.PUT and L.HPC was positively correlated with MMSE score, suggesting that lower overall excitation in these three regions was associated with lower MMSE score in MCI/AD. If the total E-I balance was assessed as E/I ratio, only one region (L.HPC) exhibited significant (positive) correlation with MMSE score (result not shown). In contrast, no significant correlation was able to pass multiple comparison correction for the association between inter-regional E-I balance and MMSE score. Taken together, regional E-I imbalance is a meaningful physiological substrate for cognitive impairment. Notably, intra-regional E-I balance as measured by local recurrent excitation and inhibition strengths shows the strongest correlation with cognitive performance, suggesting the importance of measuring and modulating intra-regional E-I imbalance for AD diagnosis and treatment. Moreover, the left HPC exhibits the most robust association with MMSE score which remains highly significant for both intra-regional and total E-I balance, and for evaluation using both E-I difference and E/I ratio. For the other six brain regions with significant correlation with MMSE score, L.PCU and L.PCC belong to the DMN, R.cACC belongs to the salience network, and L.PAL, L.PUT and R.PUT belong to the limbic network. Of note, these regions also show significant differences between NC and MCI/AD in terms of intra-regional E-I balance (Fig. 4) and total E-I balance (Fig. 12).

**Figure 15.**
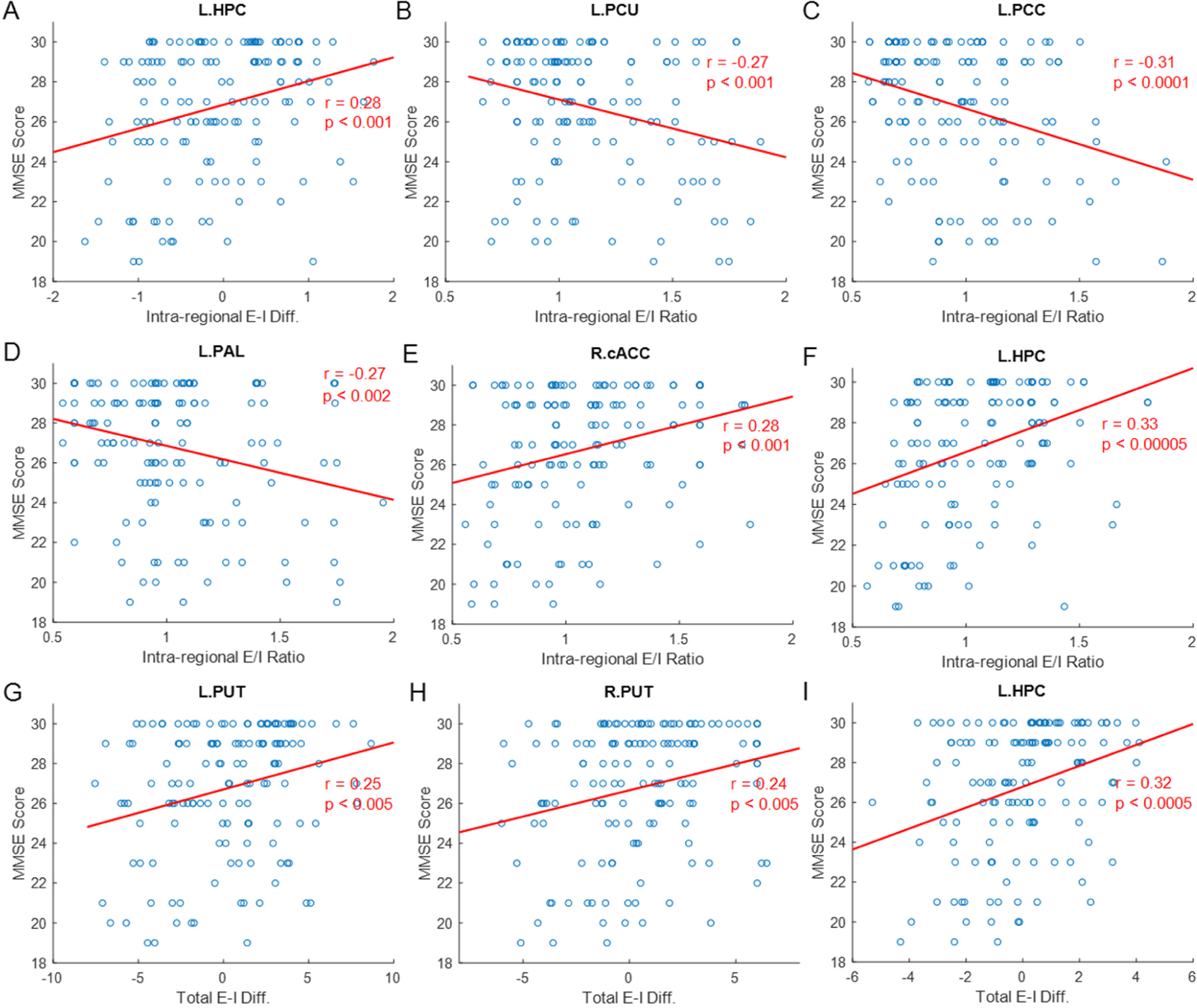
Correlation between regional E-I balance and MMSE score. **(A)** Scatter plot between intra-regional E-I difference and MMSE score. **(B-F)** Scatter plots between intra-regional E/I ratio and MMSE score for L.PCU, L.PCC, L.PAL, R.cACC, and L.HPC. **(G-I)** Scatter plots between total E-I difference and MMSE score for L.PUT, R.PUT, and L.HPC. The significance of all correlations passes FDR correction for multiple comparison (*p* < 0.05).

## Discussion

Converging evidence suggests that E-I imbalance is a critical regulator of AD pathology (Palop and Mucke, 2010; Busche and Konnerth, 2016; Palop and Mucke, 2016; Frere and Slutsky, 2018; Styr and Slutsky, 2018; Ambrad et al., 2019) and may represent a core element that underpins a “central feature” of AD by integrating pathophysiological findings from multi-levels of analysis (cell-circuit-network) (Maestú et al., 2021). Identifying pathological E-I balance during the progression of AD thus constitutes an important first step to developing new diagnostic techniques that use E-I imbalance as a biomarker and new treatment paradigms that aim to restore E-I balance. Our study provides both a novel framework to measure pathological E-I balance and important insights into the systematic features and circuit mechanisms of E-I alterations during the progression of AD.

### A multiscale neural modeling framework for E-I estimation

Due to the inability of conventional fMRI to resolve excitatory versus inhibitory activities (Devor et al., 2007; Anenberg et al., 2015; Vazquez et al., 2018), a fMRI-based computational framework that could accurately estimate E-I imbalance during AD progression is urgently needed. As the two widely used approaches for generative modeling, DCM (Friston et al., 2003, 2014; Li et al., 2011) and BNM (Honey et al., 2007, 2009; Deco and Jirsa, 2012; Deco et al., 2013a, b) are limited in either the biophysical realism (DCM) or the ability to estimate individual connection strengths (BNM) (see review in Li and Yap, 2022). To overcome these limitations, we applied a recently developed MNMI framework (Li et al., 2019; 2021) to an ADNI dataset to identify disrupted E-I balance in a large network during AD progression. Results show that MNMI is capable of identifying impaired excitatory and inhibitory EC in MCI and AD, which can be harnessed to infer E-I imbalance at a mesoscale circuit level. The accuracy and reliability of MNMI are supported by the following observations: **(1)** E-I balance is more significantly impaired in AD than MCI, consistent with the more advanced disease stage of AD; **(2)** The brain regions that exhibit the most consistent and robust E-I alterations in MCI and AD (e.g., HCP and ACC) concur with their critical roles in AD pathophysiology (see below); and **(3)** E-I imbalance in multiple brain regions is found to be significantly correlated with cognitive impairments in MCI/AD, indicating that MNMI-derived E-I alterations are behaviorally meaningful and relevant. Thus, MNMI provides a promising new tool to identify E-I imbalance in AD based on rs-fMRI.

### Systematic features of E-I alterations

One important hallmark of AD pathology is the progressive disruption of synaptic transmission (Sheng et al., 2012; Marsh and Alifragis, 2018). Consistently, we demonstrated that both excitatory and inhibitory interactions are substantially altered during the progress of AD and such alterations exhibit systematic features. <u>First</u>, excitatory and inhibitory connections are progressively disrupted during AD progression. For both intra-regional and inter-regional neural interactions, more connections are impaired in AD than MCI (when compared with NC) and the degree of impairments also becomes more significant in AD (Figs. 3, 8). As a result, E-I balance is also progressively impaired (Figs. 4, 9, 12), which is confirmed by linear model analysis (Figs. 5, 10, 13). <u>Second</u>, AD pathology differentially alters excitatory and inhibitory connections. Compared with recurrent excitatory connections, more recurrent inhibitory connections are impaired and to a greater extent, in agreement with the emerging viewpoint of GABAergic dysfunction in AD (Li et al., 2016; Palop and Mucke, 2016; Xu et al., 2020). Importantly, alterations of inhibitory connections exhibit a more stable pattern than excitatory connections as consistent impairments are observed across MCI and AD (Fig. 3). <u>Lastly</u>, AD progression is associated with a general decoupling of excitatory and inhibitory interactions. Although the strength of excitatory and inhibitory connections could either increase of decrease in MCI/AD, a reduction of connection strength dominates for both intra-regional and inter-regional connections (Table 2, Figs. 3, 8 and 11), consistent with the “synaptic dismantling” theory of AD (Selkoe et al., 2002). The heterogenous but decrease-dominated alterations in excitatory and inhibitory coupling strengths (i.e., effective connectivity) also concord with the observed bidirectional changes yet widespread decrease in functional connectivity in MCI and AD (Filippi and Agosta, 2011; Brier et al., 2014; Dennis and Thompson, 2014).

### Heterogenous alteration of E-I balance

One important finding of this study is that we observed heterogenous, region-specific alteration of E-I balance. Depending on the specific modulation of excitatory and inhibitory connections, E-I difference can either increase or decrease for different brain regions. Our findings are consistent with experimental data that hyperactive neurons coexist with hypoactive neurons in an AD mouse model (Busche et al., 2008) and MCI and AD are associated with both regional hyperactivation and hypoactivation in human (Celon et al., 2006; Corriveau-Lecavalier et al., 2019). We further revealed that increase of E-I difference is mostly due to a decrease of inhibitory connection strength (Figs. 3, 4), in agreement with experimental findings that neuronal hyperactivity is a result of decreased synaptic inhibition (Busche et al., 2008). Of note, studies have revealed that alteration of E-I balance depends on the stage of AD progression, where the HPC shows hyperactivity in early-stage aMCI, but reduced activity in late aMCI and AD (Dickerson et al., 2004, 2005; Celone et al., 2006). By comparison, our modeling results indicate that alterations in E-I balance remain consistent across MCI and AD for the same region and the (left) HCP exhibits reduced E-I difference throughout (Figs. 4, 9, 12). This may be due to the fact that elevated excitation in HPC is a temporal event in the early aMCI stage, similar to the transient increase of FC in the DMN and salience networks at the very mild AD phase (Brier et al., 2012).

### A core network of E-I imbalance in AD

Despite the heterogeneous and distributed changes in E-I interactions, we observed consistent patterns of E-I disruptions in a set of brain regions including the HPC, pallidum, putamen, nucleus accumbens, inferior temporal cortex (ITC) and caudal anterior cingulate cortex (cACC). These brain regions were consistently impaired across MCI and AD for intra-regional E-I balance (Fig. 4), inter-regional E-I balance (Fig. 9) or overall E-I balance (Fig. 12) in our study. Such a core network highlights the paramount importance of the limbic/subcortical regions and cingulate areas in AD pathophysiology. The involvement of HPC, the core region in the memory network, is consistent with the vast majority of literature about the central role of this critical structure in AD (Dickerson et al., 2004, 2005; Wang et al., 2006; Palop et al., 2007; Bakker et al., 2012). The reduction of E-I difference in HPC due to increased inhibition is also consistent with the experimental findings that high GABA content in reactive astrocytes of the dentate gyrus was discovered in brain samples from human AD patients as well as AD mouse model resulting in increased tonic inhibition and memory deficit (Wu et al., 2014). The stable participation of the basal ganglia including pallidum, putamen and nucleus accumbens in E-I disruption is somewhat unexpected as the primary function of these subcortical structures is motor control (Groenewegen, 2003). However, recent MRI studies have consistently revealed substantial volume reduction in the basal ganglia of Alzheimer’s patients, including the putamen and caudate nucleus (Cho et al., 2014; de Jong et al., 2008, 2011). The striatum, consisting of the putamen, nucleus accumbens and caudate nucleus, is particularly susceptible to AD degeneration since both Aβ plaques and neurofibrillary tangles (NFT) of hyperphosphorylated tau are found in striatal regions (Vitanova et al., 2019) and Aβ deposition starts in the striatum of presenilin-1 mutation carriers (Klunk et al., 2007). Importantly, Aβ may begin to develop in the striatum 10 years before expected symptom onset (Bateman et al., 2012), suggesting that the basal ganglia could be an important pathophysiological target in AD.

The ITC plays a critical role in visual perception, object recognition, and semantic memory processing (Ishai et al., 1999; Herath et al., 2001; Onitsuka et al., 2004). Functional deficits in these cognitive processes have been well documented in patients with MCI and AD (Hof and Bouras, 1991; Giffard et al., 2001; Laatu et al., 2003; Uhlhaas et al., 2008). It was observed that inferior temporal tau is associated with daily functional impairment in AD (Halawa et al., 2019). Disruption of E-I balance in ITC is consistent with the significant synaptic loss in this region in individuals with aMCI (Scheff et al., 2011), which may underlie early AD symptomatology. Lastly, the anterior cingulate cortex (ACC) plays a vital role in multiple cognitive processes including executive function, memory and emotion (Carter et al., 1999; Bush et al., 2000; Weible et al., 2013). It is one of the earliest affected areas by Aβ accumulation (Braak et al., 1991; Raj et al., 2012) and exhibits disrupted FC in MCI and AD (Liang et al., 2015; Liu et al., 2017). It has been demonstrated that Aβ alters E-I balance in ACC through inhibiting presynaptic GABA-release from fast-spiking interneurons onto pyramidal cells (Ren et al., 2018).

In addition to the above core AD network regions, the precuneus and PCC, the two central nodes in the DMN, show increased E-I difference in AD (Figs. 4, 9, 12) and their intra-regional E/I ratio is significantly correlated with cognitive performance as measured by the MMSE score (Fig. 15). As the main connectivity hub of DMN, the precuneus/PCC is involved in high-order cognitive functions such as emotion, arousal, self-consciousness, memory, and visuospatial processing (Maddock et al., 2003; Lou et al., 2004; Cavanna and Trimble, 2006; Wallentin et al., 2006; Leech and Sharp, 2014). It is one of the most salient areas of tau deposition and neuroinflammation (Veitch et al., 2019). Recent studies have indicated that involvement of the precuneus/PCC is significant for the development of AD (Yokoi et al., 2018) and magnetic stimulation of the precuneus was shown to slow down cognitive and functional decline (Koch et al., 2022). Increased excitation in precuneus/PCC is consistent with task-induced deactivation deficits in DMN, a robust functional impairment in MCI and AD (Lustig et al., 2003; Greicius et al., 2004; Rombouts et al., 2005). Hence, the precuneus and PCC should be considered as extended components of the core AD network whose E-I imbalance underlies key pathological changes in AD.

### Model limitations

One notable limitation of the current study is that we used DTI data from healthy HCP subjects (instead of NC/MCI/AD participants) to calculate SC, following the practice of a previous MNMI study (Li et al., 2021). Nevertheless, the impact of this limitation should be minimal since MNMI uses the average SC (from 100 HCP subjects) as a common base to constrain EC estimation (i.e., the EC is scaled by the SC which is the same for all subjects; Eqn. (1)). This average SC could mitigate the individual differences between HCP and NC/MCI/AD subjects. Also, the optimization of each individual EC parameter (*W*_kj_ in Eqn. (1)) can compensate for the potential SC difference between HCP and NC/MCI/AD subjects. Also, to remove false positive SC links and avoid over-parameterization, we estimated only 10% of the strongest inter-regional connections based on SC. We did not estimate more than 10% since it will substantially increase the number of free parameters in this large network which would reduce the estimation accuracy due to potential overfitting. Future improvement of MNMI may allow for the estimation of more EC parameters in a large-scale network.

## Conclusions

Using a multiscale neural model inversion framework, we identified disrupted regional E-I balance as well as impaired excitatory and inhibitory neural interactions during AD progression. We observed that E-I balance is progressively disrupted from NC to MCI and to AD and alteration of E-I balance varies from region to region. Also, we found that inhibitory connections are more significantly impaired than excitatory connections and the strength of the majority of connections reduces in MCI and AD, leading to gradual decoupling of neural populations. Moreover, we revealed a core AD network comprised mainly of limbic and cingulate regions exhibit consistent and stable E-I alterations across MCI and AD, which may represent an early AD biomarker and an important therapeutic target to restore pathological E-I balance. Furthermore, we found that alterations in regional E-I balance of the extended core AD network including the precuneus/PCC is behaviorally relevant by showing a significant correlation with the MMSE score.

## Methods

### Overview of MNMI

The schematic diagram of the MNMI framework is depicted in Fig. 1. The neural activity (*x*) is generated by a neural mass network model (Wilson and Cowan, 1972) consisting of multiple brain regions (R1, R2, etc.). Each region contains one excitatory (*E*) and one inhibitory (*I*) neural populations with reciprocal connections and receives spontaneous input (*u*). Different brain regions are connected via long-range fibers whose baseline strengths are determined by SC from diffusion MRI; the weak inter-regional connections are removed to avoid over-parameterization and superficial links (Li et al., 2021). The neural activity (*x*) is converted to simulated BOLD signal (*y*) via convolution with a hemodynamic response function (HRF, Friston et al., 1998) and simulated FC is computed. Both intra-regional recurrent excitation (*W*_EE_) and inhibition (*W*_IE_) weights and inter-regional connection strengths (*W*_12_, *W*_21_, etc.) as well as the spontaneous input (*u*) are estimated using genetic algorithm, a biologically inspired method for solving optimization problems based on natural selection (Mitchell, 1995), to minimize the difference between simulated and empirical FC.

### Subjects

The rs-fMRI data was obtained from the ADNI dataset (http://adni.loni.usc.edu/). A total of 144 subjects with Mini-Mental State Examination (MMSE) scores were selected from the ADNI-Go and ADNI-2 studies, including 48 normal control (NC) (26/22 males/females, age 73.4 ± 6.5 years, MMSE 29.2 ± 1.1), 48 MCI (27/21 males/females, age 73.9 ± 10 years, MMSE 28 ± 1.7) and 48 AD subjects (27/21 males/females, age 73.6 ± 8.6 years, MMSE 23.1 ± 2.5). All subjects were matched in terms of age (*p* = 0.95, one-way Analysis of Variance (ANOVA)) and gender (*p* = 0.55, one-way ANOVA).

### Image preprocessing

Data quality control was implemented in ADNI to ensure consistency across imaging centers in terms of the scanner, imaging protocol, and signal-to-noise ratio (Jack Jr et al., 2008). The fMRI data (7 min, 140 volumes) was preprocessed using AFNI (Cox, 1997) according to a well-accepted pipeline (Yan and Zang, 2010), which includes first ten volumes removal, head motion correction, normalization, nuisance signals regression, detrend and bandpass filtering (0.01 to 0.08 Hz). Nuisance regressors include head motion parameters (the “Friston-24” model), the mean BOLD signal of the white matter, and cerebrospinal fluid. To minimize artifacts due to excessive motion, the subjects with an average frame displacement (FD) (Power et al., 2014) greater than 0.5 mm will be removed. Finally, fMRI data will be smoothed with 6 mm full width at half maximum (FWHM) Gaussian kernel and then nonlinearly registered to the Montreal Neurological Institutes (MNI) space.

### Functional and structural connectivity

Regional averaged BOLD rs-fMRI time series were extracted using the Desikan-Killiany (DK) atlas (Desikan et al., 2006) with 84 regions of interest (ROIs). To reduce computational burden and focus on the networks that are most affected in AD (Zott et al., 2018), we selected 46 ROIs from the DMN, salience, executive control (frontoparietal control) and limbic networks (Table 1) based on Yeo’s seven network definition (Yeo et al., 2011) and computed the individual FC matrix using Pearson’s correlation. Structural connectivity was computed using probabilistic tractography based on the diffusion MRI data consisting of 100 unrelated subjects from the WU-Minn Human Connectome Project (HCP) young healthy adults, 1200 subjects release (Van Essen et al., 2013). The diffusion MRI data was preprocessed using the HCP protocol (Glasser et al., 2013). To compute SC, we conducted whole-brain tractography using asymmetry spectrum imaging (ASI) fiber tracking which fits a mixture of asymmetric fiber orientation distribution functions (AFODFs) to the diffusion signal (Wu et al., 2019, 2020). White matter streamlines were generated by successively following local directions determined from the AFODFs. The output streamlines were cropped at the grey/white-matter interface with a search distance of 2 mm, where the DK atlas was applied to obtain 84×84 SC matrix. The reduced SC matrix with 46 ROIs was extracted from the full SC matrix and averaged among the 100 subjects followed by normalization so the SC was bounded between 0 and 1. Finally, we selected the strongest 10% SC connections for network modeling and the weaker connections were removed (Frässle et al., 2017; Li et al., 2021).

### Neural mass model and hemodynamic response

We employed computational neuronal modeling to capture the neural interactions and dynamics in the AD network. Regional brain dynamics are simulated by a neural mass model using the biologically motivated nonlinear Wilson-Cowan oscillator (Wilson and Cowan, 1972). The population-level activity of the *j*^th^ region is governed by the following equations (Abeysuriya et al., 2018; Li et al., 2011):

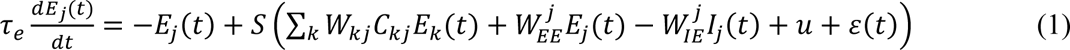

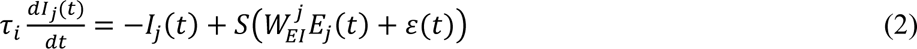

where *E*_j_ and *I*_j_ are the mean firing rates of excitatory and inhibitory neural populations in brain region *j*, *τ*_e_ and *τ*_i_ are the excitatory and inhibitory time constants (20 ms; Hellyer et al., 2016), and 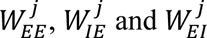 are the local coupling strengths (i.e., recurrent excitation, recurrent inhibition and excitatory to inhibitory weight). The variable *u* is a constant spontaneous input and *ɛ*(*t*) is random additive noise following a normal distribution (Deco et al., 2013a; Wang et al., 2019). The long-range connectivity strength from region *k* to region *j* is represented by *W*_kj_ which is scaled by empirical SC (*C_kj_*), and the nonlinear response function *S* is a sigmoid function 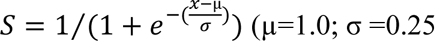; Abeysuriya et al., 2018).

To increase computational efficiency, we replaced the hemodynamic state equations in the original MNMI model (Li et al., 2021) with the canonical HRF and computed the hemodynamic response as the convolution of regional neural activity and the HRF kernel (Friston et al., 1998):

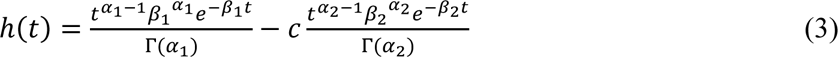

where *t* indicates time, *a*_1_ = 6, *a*_2_ = 16, *β*_1_ = *β*_2_ = 1, *c* = 1/6, and Γ represents the gamma function. The regional neural activity is calculated as the weighted sum of excitatory and inhibitory neural activity (i.e., *x*_j_ = 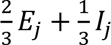; Becker et al., 2015; Li et al, 2021).

### Estimation of model parameters

Both local (intra-regional) and long-range (inter-regional) connection strengths in the model are estimated. For the local parameters, we estimated both recurrent excitation (*W*_EE_) and recurrent inhibition (*W*_IE_) weights in each ROI, resulting in 92 local parameters. The E→I coupling weight (*W*_EI_) was assumed to be constant (3.0; Li et al., 2021) as the effect of *W*_EI_ could be accommodated by change in *W*_IE_. To avoid over-parameterization and false positive connections due to DTI noise (Maier-Hein et al., 2017), we estimated the strongest 10% inter-regional connections (N = 212) and removed the remaining weaker connections. In addition, the spontaneous input (*u*) is estimated, which results in a total of 305 free parameters for estimation.

We used the genetic algorithm (GA; implemented by the *ga* function in MATLAB global optimization toolbox) to estimate the model parameters. The parameters are bounded within certain ranges to achieve balanced excitation and inhibition in the network (Li et al., 2021): *W*_EE_ and *W*_IE_ ∈ [2, 4], *W*_ki_ ∈ [-2, 2], and *u* ∈ [0.2, 0.4]. GA maximizes the Pearson’s correlation between the simulated and empirical FC matrices with the functional tolerance set to be 1e-3 and the maximal number of generations set to be 128. We observed good convergence within 128 generations for all the subjects.

### Numerical integration

The differential equations of the neural mass model are simulated using the 4th order Runge-Kutta (RK) scheme with an integration step of 10 ms; a shorter integration step has no significant effect on the results reported. We simulated the network for a total of 200 sec, and the first 20 sec of the BOLD activity is discarded to remove transient effects. The remaining 180 sec time series are downsampled to 0.33 Hz to have the same temporal resolution as the empirical BOLD signal (TR = 3 sec). The model along with the optimization procedure are coded in MATLAB and run in parallel with 24 cores in a high-performance UNC Linux computing cluster. The computing time (for each individual subject) ranges from 30 to 60 hours.

### Metrics for E-I balance

Regional E-I balance is quantified by either E-I difference (sum of incoming excitatory EC – sum of incoming inhibitory EC) or E/I ratio (the ratio of the sum of incoming excitatory EC to the sum of incoming inhibitory EC). We defined three metrics of regional E-I balance: **(1)** intra-regional E-I balance; **(2)** inter-regional E-I balance; and **(3)** total E-I balance. The intra-regional E-I balance of each region is calculated as the difference (or ratio) between recurrent excitation and recurrent inhibition strength, while inter-regional E-I balance is computed as the difference between the sum of the incoming excitatory inter-regional EC and the sum of the incoming inhibitory inter-regional EC. We did not compute inter-regional E/I ratio because some regions receive excitatory or inhibitory EC only. The total E-I balance is calculated as the difference (or ratio) between total excitation level (recurrent excitation strength + all incoming excitatory inter-regional EC) and total inhibition level (recurrent inhibition strength + all incoming inhibitory inter-regional EC) to a particular region.

## Statistical analysis

Model parameters are estimated for each subject and compared between NC and MCI, NC and AD, and MCI and AD. We used two-sample *t*-tests to compare local and inter-regional connection strengths as well as intra-regional, inter-regional and total E-I balance. Multiple comparisons are corrected by the false discovery rate (FDR; Benjamini and Yekutieli, 2001) method except for the inter-regional EC which is corrected by the Network-based Statistics (NBS; Zalesky et al., 2010) approach, both at a significance level of *p* < 0.05.

## Data availability

Both the ANDI data (https://adni.loni.usc.edu/) and HCP data (https://www.humanconnectome.org/) are publicly available. All structural and functional connectivity matrices along with BOLD fMRI time series are available from the GitHub repository (GITHUB_LINK).

## Code availability

The MATLAB codes that support the findings of this study are available from the GitHub repository given above.

## Data Availability

All data produced in the present study are available upon reasonable request to the authors.

https://adni.loni.usc.edu/

## Acknowledgements

This work was supported in part by the United States National Institutes of Health (NIH) (grant nos. EB008374, MH125479, and EB006733).

## Author contributions

G.L. and P-T.Y. conceived and designed the study. G.L. developed the model, analyzed the data, interpreted the results and prepared the manuscript. L-M.H. and Y.W. conducted data acquisition and preprocessing. G.L., A.C.B., Y-Y.I.S. and P-T.Y. discussed and revised the manuscript.

## Competing interests

The authors declare no competing interests.

Correspondence and requests for materials should be addressed to Pew-Thian Yap.

## Notes

### Competing Interest Statement

The authors have declared no competing interest.

### Funding Statement

This work was supported in part by the United States National Institutes of Health (NIH) through grants EB008374, MH125479, and EB006733.

### Author Declarations

The study used ONLY openly available human data that were originally located at Alzheimer’s Disease Neuroimaging Initiative (ADNI) database (https://adni.loni.usc.edu/).

### Summary of Updates

In this revision, we added the comparison between MCI and AD and also the correlation analysis between E-I difference/ratio and Mini-Mental State Examination (MMSE) scores. Besides, we updated some figures, revised the Introduction and Discussion.

